# West Nile virus in Italy: history and evolving transmission patterns

**DOI:** 10.64898/2026.07.12.26357540

**Authors:** Marta Giovanetti, Eleonora Cella, Vagner Fonseca, Monika Moir, Bas B. Oude Munnink, Claudio de Martinis, Ana Moreno, Alberto Rizzo, Davide Mileto, Francesca Caccuri, Arnaldo Caruso, Enzo Tramontano, Svetoslav Nanev Slavov, Ana Maria Bispo de Filippis, Tulio de Oliveira, Luiz Carlos Junior Alcantara, Alessandro Marcello, Vittorio Colizzi, Giovanni Rezza, Massimo Ciccozzi, Concetta Castilletti, Edward C. Holmes, Fabrizio Maggi, Luisa Barzon, José Lourenço

**Affiliations:** Department of Science and Bio-Technology, Università Campus Bio-Medico di Roma, Rome, Italy; Laboratório de Arbovírus e Vírus Hemorrágicos, Instituto Oswaldo Cruz, Fundação Oswaldo Cruz, Rio de Janeiro, Brazil; Burnett School of Biomedical Sciences, College of Medicine, University of Central Florida, Orlando, FL 32827, USA; Departamento de Ciencias Exatas e da Terra, Campis I, Universidade do Estado da Bahia (UNEB), Salvador, Brasil; Centre for Epidemic Response and Innovation (CERI), School fo Data Science and Computational Thinking, Stellenbosch University, Stellenbosch 7600, South Africa; Department of Viroscience, Erasmus University Medical Center, Rotterdam, the Netherlands; Department of Animal Health, Istituto Zooprofilattico Sperimentale del Mezzogiorno, Via Salute, 2, Portici, Naples, 80055, Italy; Virology Department, Istituto Zooprofilattico Sperimentale della Lombardia e dell’Emilia Romagna; Laboratory of Clinical Microbiology, Virology and Bioemergencies, Luigi Sacco Hospital, Regional Center for Infectious Diseases (CEREMI), Lombardy Region, Milan, Italy; Section of Microbiology, Department of Molecular and Translational Medicine, University of Brescia, Brescia, Italy; Department of Life and Environmental Science, University of Cagliari, Cagliari, Italy; Butantan Institute, São Paulo, Brazil; Rene RachouInstitute, Oswaldo Cruz Foundation, Belo Horizonte, Brazil; International Centre for Genetic Engineering and Biotechnology (ICGEB), Trieste, Italy; Faculty of Medicine, University Hospital Complex “Le Bon Samaritain”, N’Djamena, Chad; Tor Vergata University Foundation, Rome, Italy; Faculty of Medicine, Vita-Salute San Raffaele University, Milan, Italy; Unit of Medical Statistics and Molecular Epidemiology, University of Campus Bio-Medico di Roma, Rome, Italy; IRCCS Sacro Cuore Don Calabria Hospital, Negrar, Italy; School of Medical Sciences, The University of Sydney, Sydney, New South Wales, Australia; Laboratory ofVirology and Laboratories of Biosafety, National Institute for infectious Diseases “Lazzaro Spallanzani” – IRCCS, Rome, Italy; Department of Molecular Medicine, University of Padova, Padova, Italy; Universidade Católica Portuguesa, Católica Medical School, Católica Biomedical Research Centre, Rio de Mouro, Portugal

**Author notes:** **Correspondence:** Marta Giovanetti.

**Keywords:** West Nile virus, genomic surveillance, phylodynamics, climate change, Italy

## Abstract

**BACKGROUND:** West Nile virus (WNV) has become an important public health concern in Europe. Italy is one of the most affected countries, yet our understanding of WNV epidemiology, genomics, and dispersal across hosts and geographic regions is incomplete.

**AIM:** To reveal the history of WNV in Italy by integrating epidemiological, genomic, and environmental data into descriptive and quantitative assessments of its past spatio-temporal surveillance and expansion.

**METHODS:** We collated vertebrate and mosquito WNV records from national surveillance and the scientific literature spanning multiple decades. Historical serological and molecular data were summarized by host and region, climatic associations with case trends were assessed using regression models, and phylodynamic and phylogeographic analyses reconstructed viral introductions and dispersal within Italy.

**RESULTS:** WNV circulation in Italy has changed markedly over time, with increasing human case reporting and expansion beyond historically affected northern regions. Climate-informed regression models explained recent reporting trends, supporting an environmental contribution to transmission. Phylodynamic analyses identified multiple independent introductions and sustained local transmission with increasing regional connectivity. Wavefront analyses revealed lineage-specific dispersal patterns associated with seasonal climatic gradients. Discrepancies between epidemiological records and genomic sampling highlighted uneven surveillance across regions and host species.

**CONCLUSION:** WNV emergence in Italy reflects repeated viral introductions, local persistence, heterogeneous surveillance, and environmentally associated dispersal dynamics. Strengthening integrated surveillance combining epidemiological, environmental, and genomic data will improve early detection, the monitoring of transmission dynamics, and public health preparedness under ongoing environmental change.

## Introduction

West Nile virus (WNV), a mosquito-borne flavivirus maintained in enzootic transmission cycles between birds and *Culex* spp. mosquitoes, is one of the most important and persistent arthropod-borne (arboviral) threats across Europe and the Mediterranean basin [1]. Since its establishment in temperate regions, WNV has caused recurrent outbreaks of West Nile neuroinvasive disease (WND) in humans and substantial impacts on animal health, demonstrating a marked ability for persistence, amplification, mixing, and re-emergence across diverse ecological settings [1,2]. Over the past two decades, WNV activity in Europe has intensified, with increasing case notifications, progressive spatial expansion, and recurrent seasonal outbreaks [1]. These dynamics reflect a complex interplay between vector and zoonotic host ecologies, human communities, and environmental drivers, positioning WNV as a paradigmatic climate-sensitive pathogen [3-5].

Italy represents one of the primary hotspots of WNV circulation in Europe, characterised by sustained transmission and repeated epidemic activity over the past decade [6]. According to national surveillance data from the Istituto Superiore di Sanità (ISS), annual human WND cases have ranged from fewer than 10 cases in low-transmission years to over 600 cases during major outbreaks, with notable epidemic peaks in 2018 (n≈580) and 2022 (n≈600) [7]. While early circulation was largely confined to northern regions, recent patterns indicate a clear geographic expansion driven by evolving transmission dynamics at the subnational scale. Despite surveillance efforts, critical gaps remain in our understanding of the long-term patterns and drivers of WNV circulation in Italy [7]. These gaps should also be interpreted in the context of the progressive evolution of the Italian surveillance system. Over the last two decades, surveillance has expanded from fragmented host-specific activities towards an increasingly integrated One Health framework, although surveillance intensity, the hosts monitored and diagnostic strategies continue to differ across regions and host species. Consequently, historical epidemiological patterns may partly reflect changes in surveillance sensitivity in addition to changes in viral circulation.

Here, we present an integrated analysis of the historical circulation of WNV in Italy, combining epidemiological, environmental, and genomic data and analyses. Using this integrative approach, we aimed to (i) characterise the spatio-temporal structure of WNV transmission across hosts, (ii) quantify the relationship between climatic drivers and human case numbers, and (iii) reconstruct the introduction and dispersal pathways of WNV lineages within Italy. In parallel, this unified perspective reveals the role of climate change in infectious disease emergence and highlights gaps in epidemiological surveillance.

## Results

### Historical evidence of WNV circulation in Italy

We collated WNV case records (infections) from the Italian Ministry of Health covering the period 2012 to 2025 and prevalence (serology) or molecular positivity rate (RT-PCR) records from the scientific literature via a systematic literature review spanning 1998 to 2024 (see **Methods, Figure S1**). Cases, seroprevalence and positivity rate were analysed considering both their reported geographic resolution (e.g., longitude-latitude coordinates, village, town, city, etc.) and/or their reported macroregion (North-East, North-West, Centre, South, Islands).

Seroprevalence and positivity rate records (**Figures 1A, S2**) included evidence across humans, birds, equines and other vertebrates, with a 95% quantile of reported range 0.0 - 7.1% in humans, 0 - 10.1% in birds, 0 - 45% in equines and 0 - 84% in other vertebrates (**Figure S3**). The North-East accounted for the largest proportion of records (40%), followed by the Centre (19%), North-West (17%), South (15%) and Islands (8%). Only 2% of records were reported at the nationwide level, pertaining to studies describing screenings of solid organ donors and migratory birds.

**Figure 1.**
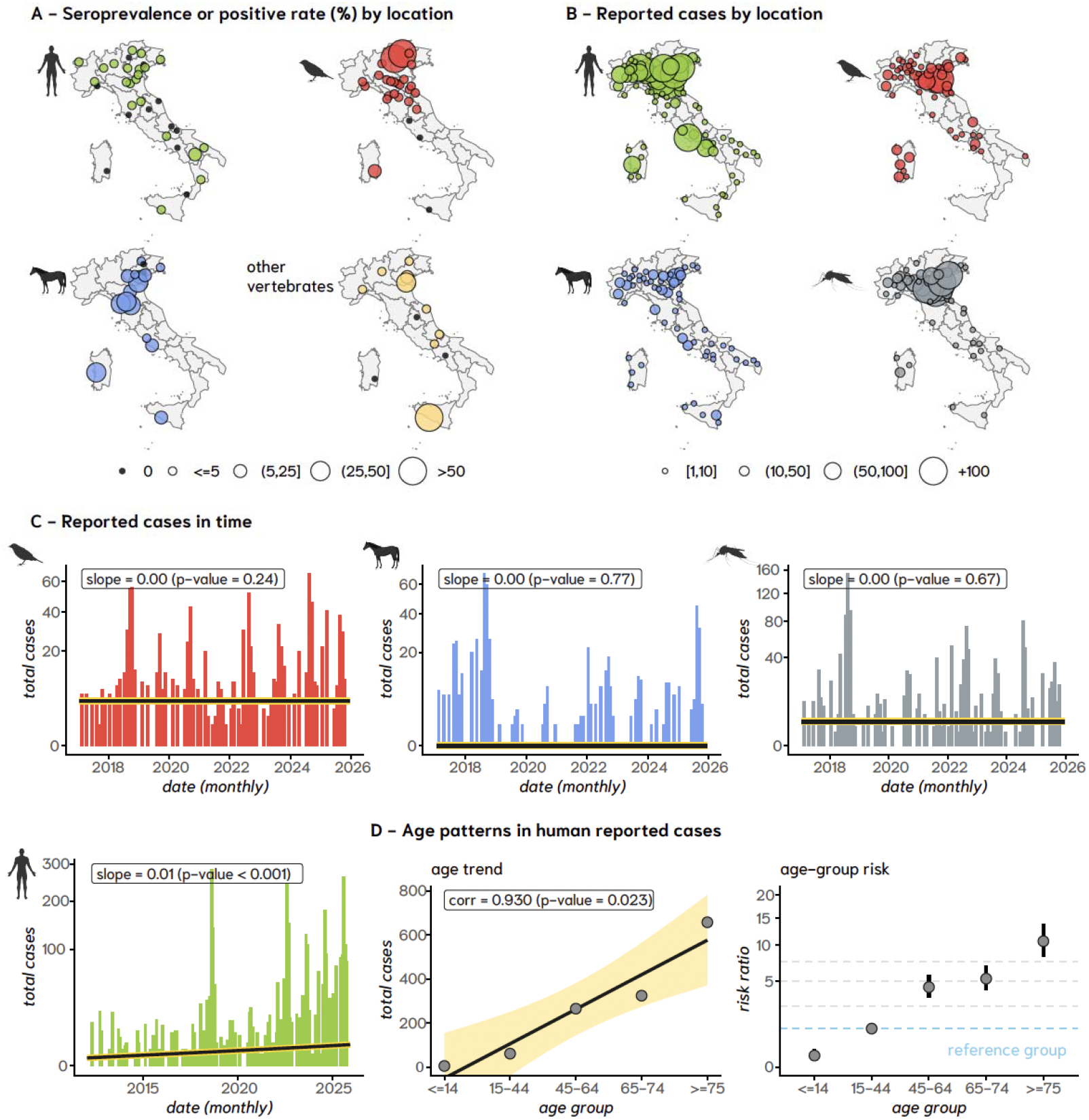
Historical evidence of WNV circulation in Italy across hosts, space, and time. A) Spatial distribution of seroprevalence (serological testing) or positive rates (molecular detection) reported in the literature, stratified by host (humans, birds, equines, and other vertebrates). Circle size reflects the magnitude of reported prevalence or positivity, with black circles marking regions for which studies reported zero. Maps intended to showcase spatial distribution and heterogeneity; (B) Spatial distribution of reported WNV cases across hosts (humans, birds, equines, and mosquitoes). Circle size represents the number of reported cases at each location, presenting solely regions with at least one case. Maps intended to showcase spatial distribution and heterogeneity; C) Temporal dynamics of reported cases (monthly) by host. Human cases show a statistically significant increasing trend over time (slope = 0.0065, p < 0.001), whereas no significant trends are observed for birds, equines, or mosquitoes. Solid lines represent fitted linear trends. The Y-axis is square-root transformed for improved visualization; D) Age distribution of reported human cases at the national level. Left: total cases by age group with corresponding trend (correlation coefficient and p-value indicated). Right: age-specific risk ratios derived from a Poisson generalized linear model, using the 15–44 age group as reference (blue dashed line; RR=1). The Y-axis is square-root transformed for better visualization.

Case records included data from humans, birds, equines and mosquitoes (**Figures 1B, S4**). Historical seasonal dynamics were consistent across hosts, with a marked peak in summer (July–September) and a trough in winter–spring (December–May) (**Figure S4**). The month with the historically lowest reporting was December, whereas the highest reporting occurred in August, followed by September and July. Across vertebrate hosts, humans accounted for the majority of historical records (66%), followed by birds (22%) and equines (12%). These host-specific differences likely reflect, at least in part, differences in surveillance intensity, monitored host populations, and reporting strategies adopted across the study period. The North-East represented the largest proportion of records (63%), followed by the North-West (21%), the Centre (7%), and the Islands and South (both 5%).

Monthly time series of case records (**Figure 1C**) showed an increase in reporting of human cases over time, whereas no trends were observed for birds, equines or mosquitoes. The historical slope of monthly median human case reporting was approximately 0.0065 (p-value<0.001), corresponding to an 8.1% year-on-year median increase of monthly reporting between 2012 and 2025. There was also evidence of underlying spatio-temporal expansion trends in human records. For example, there was an increase of one more macroregion with at least one record every 6.6 years (slope=0.15, p-value=0.05) (**Figure S5**). This trend varied seasonally, with an increase in detections every 10 years in winter (p-value=0.016), 7 years in autumn (p-value=0.042), 4.2 years in spring (p-value=0.024) and 3.7 years in summer (p-value=0.005) (**Figure S5**). At the same time, the number of months with case detections significantly increased every 3 years (p-value=0.01) (**Figure S5**).

Nationwide, there was a positive correlation of 0.93 (p-value=0.023) between total human case records and reported age-groups (**Figure 1D**). The mean age of reporting was estimated to the midpoint of age-groups via two methodologies (see **Methods**), resulting in similar estimates of ~70.6 (~69.9 - 71.4, 95% CI) for both bootstrap and weighted mean approaches (**Figure S6**). Using the age-group 15-44 years as reference, the risk-ratio of reporting was 0.09 (0.04 - 0.21, 95% CI) for the group <=14 years, 4.32 (3.3 - 5.7) for the group 45-64 years, and 10.7 (8.2 - 13.9) for the group >=75 years (**Figure 1D**). In effect, without population size normalization, the risk of reporting in individuals aged >=75 years was approximately 119-fold higher than in those under 15 years. The positive correlation between cases and age was also found per macroregion, leading to macroregion-specific estimates of mean age and risk-ratio of reporting that were very similar to those at the nationwide scale (**Figure S6**).

### Long-term trends in climate and WNV human reporting

Given the well-established role of local climate in shaping WNV transmission potential [8] and the statistically significant increase in human case reporting observed in Italy between 2012 and 2015 (**Figure 1C**), we compared historical climate patterns at both the nationwide and macroregional levels with annual case records. Between 1979 and 2025, Italy experienced a general trend toward a warmer and dryer climate, characterized by a statistically significant increase in temperature (**Figure 2A**), a decrease in humidity (**Figure 2A**) and stable precipitation across both national and macroregional scales (**Figure S7**). During the period 2012 to 2025, human case records were positively correlated with temperature at a nationwide scale (0.8, p-value<0.001; **Figure 2B**) and within the North-West macroregion (0.75, p-value=0.003; **Figure S8**). In contrast, human case records were not correlated with humidity at the nationwide (**Figure 2B**) or macroregion levels (**Figure S9**). During the same time period, mosquito records were not statistically correlated with either temperature (0.5, p-value=0.17) or humidity (0.45, p-value=0.23) at a nationwide scale. The lack of a significant association between climate variables and mosquito positivity may partly reflect the design of the Italian surveillance system, where entomological sampling is largely restricted to the seasonal period when climatic conditions are already favourable for WNV transmission, thereby reducing the environmental variability captured by surveillance.

**Figure 2.**
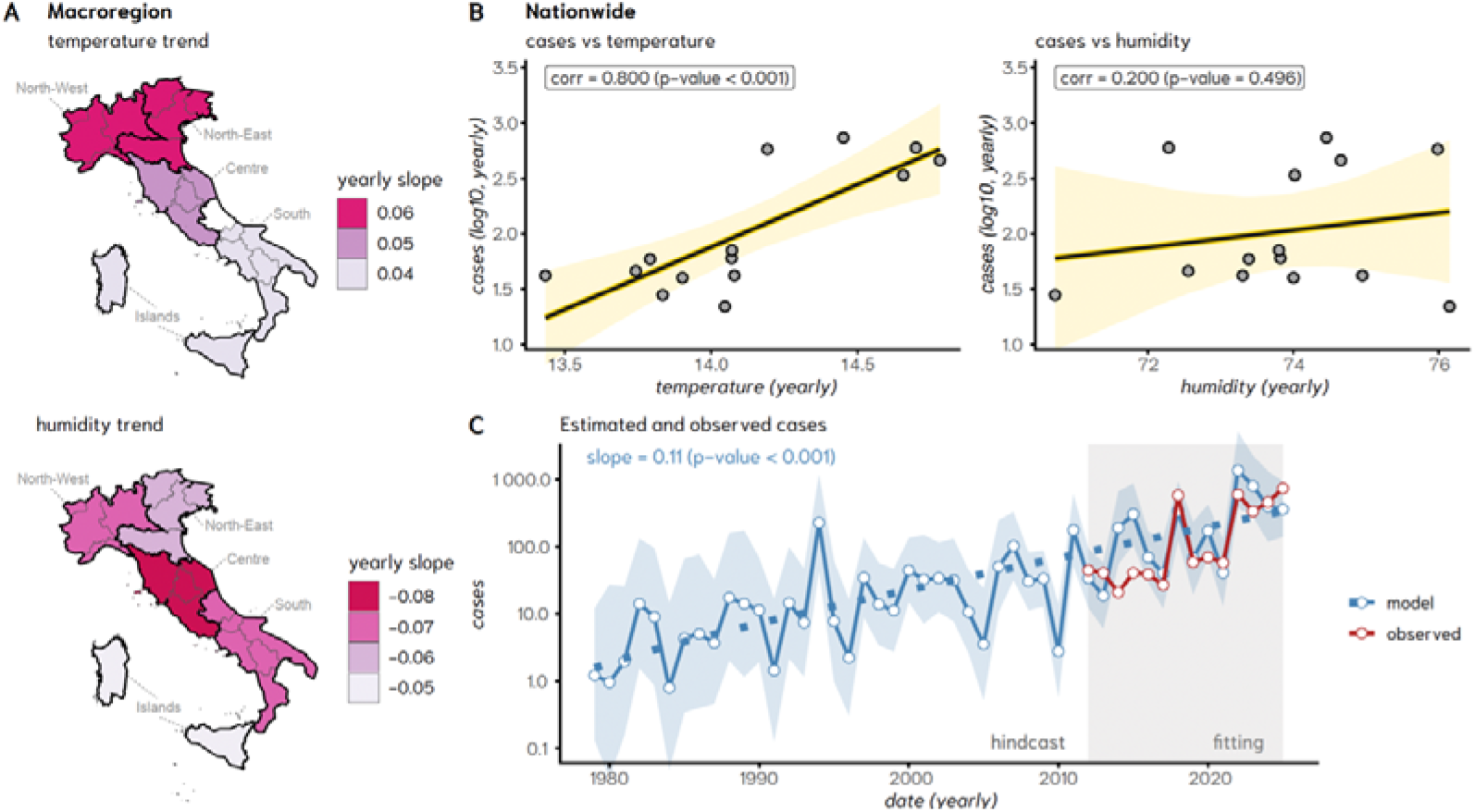
Long-term climatic trends and their relationship with reported human cases of WNV in Italy. A) Spatial distribution of climatological trends (median slope year-on-year) across Italian macroregions for temperature (top) and humidity (bottom). B) Nationwide relationships between yearly climate summaries (temperature (°C)-left; humidity (%) - right) and total reported human cases (log_10_-transformed). C) nbGLM output (mean - blue line; shaded blue - 95% CI) as informed by historical climate data from 1979 to 2025 (temperature, humidity, and precipitation) and observed human cases (red). The grey background area highlights the fitting time window, and the white background area the hindcast time window.

Annual human case records in the reporting period 2012 to 2025 were modelled using a negative binomial Generalized Linear Model (nbGLM) at the macroregion level with temperature, humidity, and precipitation as predictors, and the macroregion Center as reference (see **Methods** for details and extra outputs). The nbGLM explained a substantial proportion of deviance relative to a null model (McFadden pseudo-R^2^ = 0.49) with all climatic predictors and macroregion fixed effects reaching statistical significance. As reflected in nbGLM coefficients (see **Methods**) and predictor partials (**Figure S10**), temperature exerted the strongest positive influence on case records, followed by humidity, while precipitation had a significant negative effect. Correlation of model fitted values versus observed records was highest in the North-West and North-East (~0.7, **Figure S10**), which are the two macroregions accounting for the majority of case records (**Figure S4**). In contrast, the correlation was intermediate in the Islands (0.44) and low for the South (0.28) and Centre (0.20) (**Figure S10**), the latter two being characterised by a high frequency of zero-count years (**Figure S4**). Despite the variation in correlations between the nbGLM fitted and observed case records across the macroregions, when predictions were aggregated to the national level (**Figure 2C**), nbGLM performance was adequate. Indeed, the nationwide root mean squared error (RMSE) and mean absolute error (MAE) were approximately 70% and 87%, respectively, of the summed macroregional errors (**Figure S10**), indicating low systematic national bias and supporting the use of macroregions as a fixed rather than random effect. Hindcast nbGLM estimations using climatic predictors between 1979 and 2025 (as available in the climate data set, see **Methods**) suggested that recent trends in nationwide human reporting (2012-2025) could be explained by a linear median year-on-year increase of ~11% since the early 1980s (**Figure 2C**), when cases would have been largely absent from Italy.

### Evolutionary and spatial dynamics of WNV in Italy

To investigate the introduction and dispersal dynamics of the WNV lineages circulating in Italy, we performed phylogeographic reconstruction integrating time-scaled phylogenies and spatial diffusion analyses of historical genome sequences (**Figure 3**).

**Figure 3.**
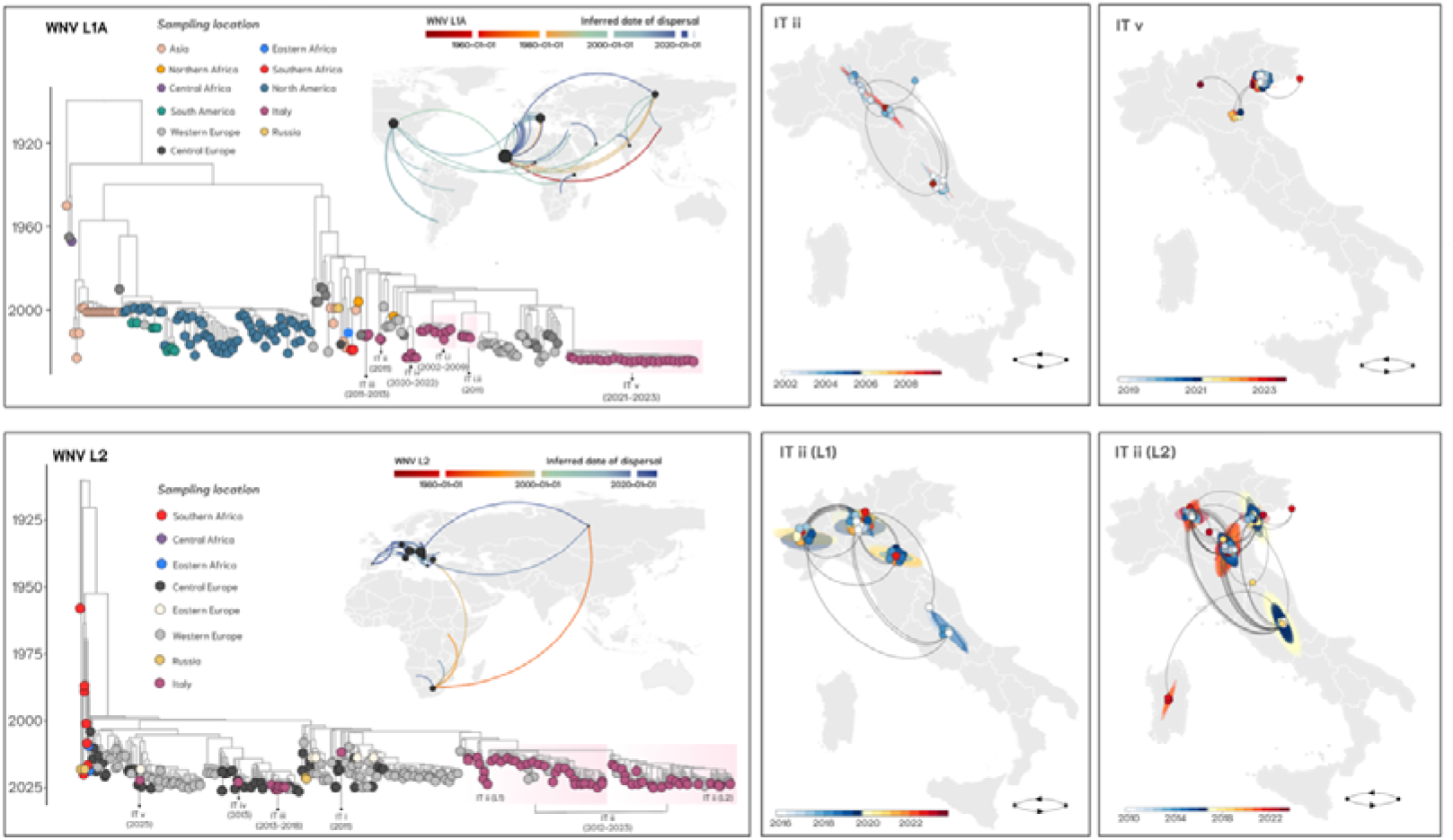
Phylogeographic reconstruction of West Nile virus (WNV) lineages in Italy. Time-scaled phylogenies of WNV lineage 1a (L1a) and lineage 2 (L2) are shown alongside inferred viral dispersal patterns. Tips are colored according to sampling location, and major clades are highlighted. For visualization purposes, non-European sampling locations are represented by country-level centroid coordinates. Spatiotemporal diffusion is represented by curved lines indicating inferred transmission routes, which are colored by the estimated date of dispersal. For WNV L1a, global dissemination patterns and introductions into Italy are illustrated, followed by regional spread within the country (clades II–IV). For WNV L2, the figure depicts its introduction and subsequent expansion across Europe and Italy, including detailed transmission patterns within northern and central regions (subclades II (L1) and II (L2)). Color gradients indicate the temporal progression of viral movement.

For lineage 1a (L1a), the phylogeny reveals multiple independent introduction events into Italy within a broader Afro-Eurasian context (**Figure 3**). Following the introduction, L1a diversified into several geographically structured clades (denoted as IT i–v). Clade L1a IT i represents early circulation and is further subdivided into clades L1a IT i.i and IT i.ii, indicating distinct introduction and establishment events during the early 2000s. Clade L1a IT ii represents a subsequent introduction and spread in Italy, followed by clades L1a IT iii and L1a IT iv, which reflect subsequent diversification and localized persistence. Clade L1a IT v represents the most recent lineage (2021–2023), forming a well-supported and temporally distinct cluster. Among these, only a subset of clades provided sufficient phylogenetic and spatial signals to support phylogeographic reconstruction. In particular, clade L1a IT ii captures early circulation dynamics, with phylogeographic reconstruction indicating inferred viral lineage movements concentrated in northern Italy, especially within the Po Valley, alongside connections with central and southern regions (**Figure 3**, right upper panels). This phase is characterized by broader geographic connectivity compared with later clades. In contrast, clade L1a IT v reflects more recent transmission dynamics, with spatial reconstruction showing highly localized viral lineage dispersal largely confined to northern Italy. Inferred dispersal events are predominantly limited to short-range exchanges between neighbouring areas, with no evidence of large-scale geographic expansion.

In contrast, lineage 2 (L2) exhibits a more phylogenetically structured pattern, with samples from South Africa, where endemic circulation is well documented, occupying basal positions in the phylogeny, consistent with its important role in the documented evolutionary history of WNV L2 (**Figure 3**). From this background, the European component forms a large geographically structured cluster, consistent with diversification across the continent. Within this framework, the phylogeny resolves the principal Italian clusters (L2 IT i to IT iv), as well as a recently emerging L2 IT v cluster. A clear subdivision is observed within IT ii, comprising two distinct components: L2 IT ii (L1) and L2 IT ii (L2). Spatial reconstructions indicate that L2 underwent a major introduction into Europe followed by rapid expansion into Italy. Within the country, L2 IT ii (L1) exhibits geographically clustered transmission patterns with inferred movement links across northern Italy, including west–east and north–south connectivity. In contrast, L2 IT ii (L2) exhibits a more extensive and complex spatial diffusion, characterized by dense transmission networks spanning northern and central Italy and clear evidence of long-distance connectivity, including links toward southern regions. Earlier clusters, including L2 IT i and L2 IT iii, exhibit more limited spatial dispersion and appear less connected within the contemporary transmission network, whereas L2 IT iv reflects more recent circulation with a relatively restricted geographic distribution. Notably, the L2 IT v cluster corresponds to a recent reintroduction event detected in 2025. Although represented by only a limited number of genomes in the present dataset, this cluster includes viruses detected in central Italy, consistent with recent genomic reports describing the emergence of WNV lineage 2 in central and southern Italy during the most recent transmission seasons [8]. At the time of analysis, however, genomic evidence remained insufficient to infer widespread dissemination. The corresponding viral movement reconstructions for L2 IT ii (L1) and L2 IT ii (L2) clades (**Figure 3**, right bottom panels) indicate frequent bidirectional exchanges and overlapping transmission pathways, particularly within L2 IT ii (L2), consistent with sustained circulation and high regional connectivity. The temporal framework inferred from the phylogenetic analyses indicates multiple independent introduction and persistence events across Italy. Detailed estimates of Times of Most Recent Common Ancestry (tMRCA) and associated 95% highest posterior density (HPD) intervals are reported in Supplementary **Table S1**.

### Viral wavefront distance dynamics and underlying climate gradients

To further investigate the spatial expansion dynamics of WNV lineages circulating in Italy, we quantified lineage-specific wavefront distance dynamics through time (**Figure 4A**). Distinct lineage-specific dispersal profiles were observed across all analyzed clades. WNV L1A clade IT ii exhibited a progressive expansion phase between 2002 and 2008, characterized by sustained increases in wavefront distance and inferred intermediate dispersal jumps, although these early long-distance patterns should be interpreted cautiously given potential sampling heterogeneity during the initial surveillance period. In contrast, clade IT v showed a highly restricted spatial footprint, being virtually absent through most of the observation period, followed by an abrupt expansion event during the most recent phase of circulation. WNV L2 (IT ii L1) displayed an early and rapid expansion phase around 2015–2016, followed by relative spatial stabilization and only limited subsequent dispersal fluctuations. Conversely, WNV L2 (IT ii L2) exhibited the largest overall spatial expansion, with major inferred long-distance dispersal events during the early emergence phase and an additional expansion period around 2018–2020, consistent with broader geographic circulation across Italy.

**Figure 4.**
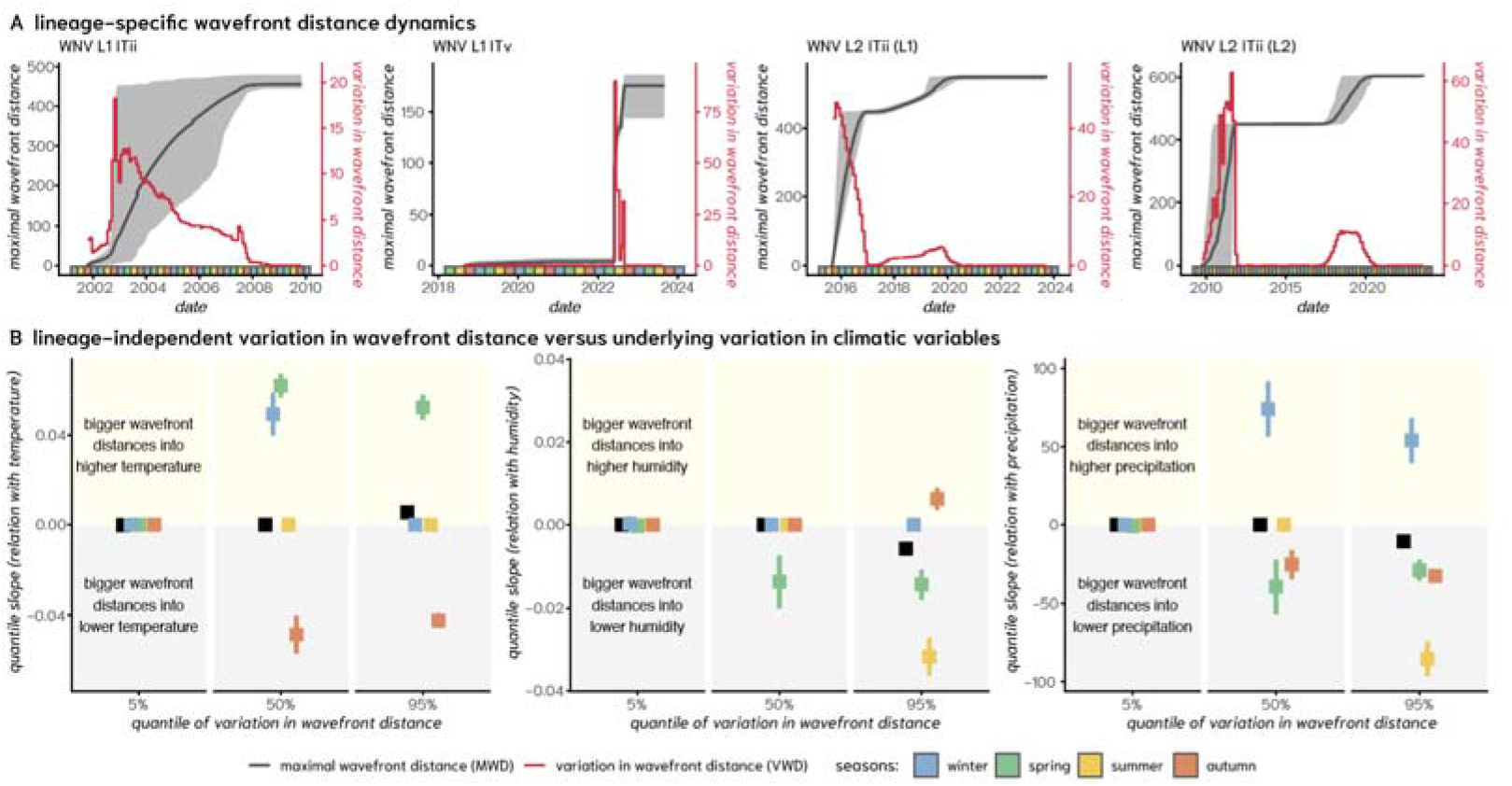
Viral wavefront distance dynamics and associated climatic gradients in WNV lineages circulating in Italy. A) Lineage-specific wavefront distance dynamics for WNV L1A clades ii, v, and for WNV L2 clades IT ii L1, and ITii L2 through time. Dark grey lines indicate maximal wavefront distance (MWD) from the inferred epidemic origin (Figure 3), with shaded areas representing the 95% HPD interval. Red lines represent temporal variation in wavefront distance (VWD), reflecting the magnitude of dispersal jumps between consecutive time intervals. Colored bars along the x-axis indicate seasonal periods (winter, spring, summer, autumn, following the color scale at the bottom); B) Quantile-based associations between variation in wavefront distance (VWD) and climatic gradients (temperature, humidity, and precipitation). Quantile slopes describe the relationship between dispersal variation and climatic change across the 5%, 50%, and 95% quantiles of wavefront variation (squares with whiskers the 95% CI). Positive values indicate movement toward regions with higher values in the respective climatic variables, whereas negative values indicate movement toward lower values. Only statistically significant associations are shown, with non-significant estimates were set to zero for visualization purposes. Colors indicate seasonal stratification, with black indicating all seasons together.

To evaluate whether these dispersal dynamics were associated with environmental gradients, we quantified the relationship between estimated variation in wavefront distance (VWD) and variation in climatic variables related to the source-sink locations connected to the dispersal jumps (**Figures S11**,**12**). When analyzed per season (**Figure S13**), the quantile-regression of variation in wavefront distance (VWD) revealed that a proportion of intermediate-to-long dispersal events showed associations with regional climatic gradients between inferred source and destination locations (**Figure 4B**). Given the limited number of inferred dispersal events, these associations should be interpreted as exploratory patterns rather than evidence of specific climatic drivers. For temperature, intermediate-distance dispersal events (quantile 50%) occurred more frequently over increasing temperature gradients in winter and spring, and over decreasing gradients in autumn. Long-distance events (quantile 95%) showed similar patterns but with the exclusion of winter. In contrast, humidity and precipitation exhibited less consistent seasonal associations with dispersal dynamics. Generally, the strongest associations with climatic gradients were observed for longer dispersal jumps (quantile 95%), suggesting that climatic suitability may have contributed to shaping historical spatial expansion patterns of WNV in Italy, potentially through effects on mosquito and avian host ecology.

### Epidemiological and genomic sampling across hosts and regions

The total number of case records was compared with the number of publicly available viral genome sequences at the macroregion level, stratified by host type (human, bird, equine, and mosquito). A statistically significant positive association between sequence availability and historical case load per macroregion was observed only for the human and mosquito hosts (**Figure S14**). In the human data, a clear geographic dichotomy emerged, with the North-East and North-West showing consistently high numbers of both cases and sequences, while other macroregions exhibited low numbers for both indicators (**Figure 5A**). In the case of birds and mosquitoes, a similar north versus centre-south spatial pattern was observed for mainland macroregions, while the Islands’ macroregion represented a notable discrepancy with limited genomic sequencing despite a relatively high number of case records. For the equine data, no macroregion showed concordant high levels of both indicators. Instead, the North displayed relatively low genomic sequencing despite high case records, whereas the South and Islands showed higher sequencing effort in the context of lower reported case numbers. This apparent mismatch may reflect the characteristics of equine WNV surveillance, where most reported cases are identified serologically, limiting opportunities for genome sequencing.

**Figure 5.**
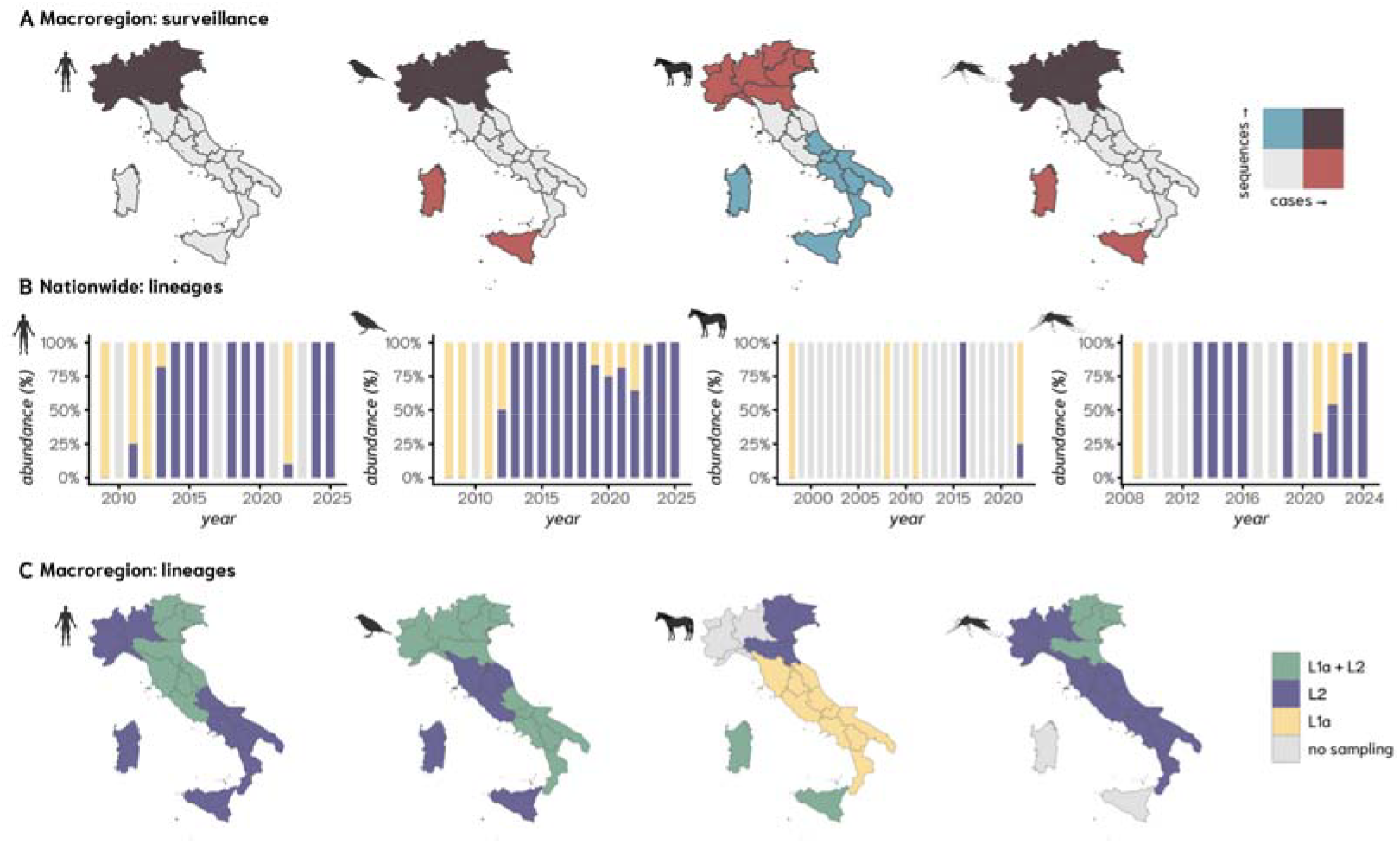
Epidemiological records and genomic sampling in time and space. A) Maps presenting the bivariate spatial relationship between (total) case records and viral sequences at a macroregion level (the midpoints of the 2×2 color scale are the median of each variable); B) Nationwide yearly lineage dynamics showing the relative temporal abundance (%) of WNV lineages (L1a - yellow, L2 - purple, no sampling - grey) in Italy over time; C) Maps presenting the spatial presence of WNV lineages at a macroregional level over all the years (L1a - yellow, L2 - purple, both - green, no sampling - grey). Significance codes: *** p < 0.001, * p < 0.05. Reference macroregion: Centre.

Historically, at the nationwide scale, between 1998 and 2025, 55% of available viral genome sequences were obtained from birds, followed by mosquitoes (23%), humans (19%), equines (2%), and other mammals (1%). Prior to 2007, genomic data were extremely limited, with only a single equine-derived sequence available (**Figure S15**). Following the implementation and subsequent expansion of the Italian national WNV surveillance programme, sequencing activity increased steadily from 2007 onwards, with a marked acceleration after 2021, particularly in mosquitoes and birds. Although human-derived genomic sampling remained limited (mean, 4.5 sequences per year), temporal lineage dynamics across host types were broadly consistent with the phylodynamic reconstruction (**Figure 3**), suggesting that WNV lineage 1a (L1a) predominated during the early phase of genomic surveillance before lineage 2 (L2) became predominant in more recent years (**Figure 5B**). The spatial distribution of L1a and L2 sampling also varied over time (**Figure 5C**; **Figure S16**). The North-East was the only macroregion with genomic sampling of both lineages across three of the four host groups. In contrast, no equine-derived genomes were available from the North-West, and no mosquito-derived genomes were available from the Islands.

## Discussion

Across Europe, West Nile virus (WNV) has progressively shifted from an episodic, emerging arbovirus to a recurrent and established one and, in some regions, to an endemic public health threat [9]. This transition reflects the convergence of environmental, avian host and vector ecologies that collectively trigger temporary or sustained transmission cycles [10,11]. Recent continental-scale analyses suggest that interannual variability in WNV outbreaks is increasingly driven by climatic anomalies, particularly temperature extremes and seasonal patterns that modulate vector-host interactions [12,13]. Consistent with these observations, our analyses demonstrate that variation in viral dispersal is closely associated with seasonal climatic gradients, further supporting the hypothesis that climate plays a key role in shaping the spatial dynamics of WNV transmission in Italy. Within this evolving landscape, Italy represents a paradigmatic setting for investigating long-term WNV transmission dynamics across temporal, spatial, environmental, host, vector, epidemiological, and genetic dimensions.

The data patterns and analyses we present strongly indicate that there is local transmission of WNV in Italy which is intensifying and reaching a broader spatial range [8]. Wavefront dynamics for each viral lineage consistently reveal a shared pattern of spatial dispersal over hundreds of kilometers within small temporal windows, indicative of rapid emergence and dispersal followed by geographically constrained transmission. Rather than reflecting isolated epidemic events, the progressive involvement of multiple macroregions suggests a transition toward broader ecological establishment within the country. This aligns with observations from other European settings where WNV has expanded beyond traditional hotspots and increasingly occupies a wider ecological niche [14,15].

The strong contribution of temperature to explain recent case reporting trends reinforces a well-established mechanistic understanding that temperature regulates mosquito life cycles, behaviour, and shortens viral extrinsic incubation periods, with the potential to both hamper and amplify transmission potential [16,17]. The contribution of humidity, although more moderate, is consistent with its role in sustaining vector survival and activity, while the negative association with precipitation likely reflects the complexity of larval habitat dynamics, where excessive rainfall can either disrupt or favour breeding sites, resulting in non-monotonic ecological effects [14]. The consistency of these observations with previous field research [4], modelling efforts across southern Europe [4,18,19], as well as with recent continent-wide analyses [20], reinforces the point that climatic drivers are not context-specific but operate as generalizable regulators on WNV transmission. The ability of climate-informed models to capture transmission patterns further highlights their potential utility for outbreak forecasting. In particular, integrating climatic projections with real-time epidemiological and genomic surveillance can support the identification of high-risk transmission periods and strengthen preparedness strategies across Europe [4, 18,19].

Beyond individual contributions of climatic variables in explaining recent case reporting, our modelling provided insights into the interpretation of long-term trends, suggesting a sustained increase in WNV case reporting over time and hence that transmission in temperate regions is undergoing gradual intensification. However, the reported constancy of this increase should be interpreted with caution, as it is partly driven by the linear assumptions supporting the modelling. In reality, WNV dynamics are inherently episodic and shaped both by stochastic and seasonal amplification cycles, meaning that the reported linear trend simply reflects a broader and likely time-varying environmental forcing. Modelling also suggested an upward trend even in macroregions with historically lower and erratic incidence, such as the Islands, where fitting performance was weaker. This discrepancy may indicate underreporting or reduced surveillance sensitivity in these regions, suggesting that WNV circulation could be more widespread than currently documented. This interpretation is consistent with previous work highlighting the risk of underestimating transmission in regions with lower surveillance sensitivity [11]. Alternatively, it suggests that environmental suitability is increasing in such regions, but that transmission is yet to become stable. In contrast, the better modelling performance in northern Italy likely reflects both greater transmission intensity and more robust surveillance systems.

The wavefront-based analyses provided additional insight into how climatic conditions may have shaped the historical spatial dynamics. Per lineage wavefront trajectories consistently showed rapid geographic expansion. Trajectories and historical spatial reach were not uniform across lineages, with some clades maintaining geographically constrained circulation while others exhibited broader regional connectivity. Notably, intermediate-to-long dispersal events were consistently associated with seasonal climatic gradients. Seasonal associations with climatic gradients varied across the year, with winter and spring generally associated with increasing temperature gradients, whereas summer and autumn showed less consistent patterns. We were unable to identify actual mechanisms of causation, but they are likely driven by host and mosquito ecologies that change by season.

Collectively, these quantitative findings suggest that WNV dispersal is not spatially random but instead follows environmentally variable corridors shaped by seasonal climatic suitability. This is consistent with previous studies demonstrating the role of climatic suitability in structuring WNV transmission dynamics and geographic expansion across Europe [8,10,18]. Our approach and outputs complement such evidence by suggesting that climatic conditions may have historically facilitated both the scale and directionality of WNV dispersal dynamics in Italy.

From an evolutionary perspective, the phylodynamic results highlighted the dual process of repeated introduction and local persistence. The presence of multiple independent introduction events was consistent with repeated long-distance dispersal across Afro-Eurasian regions, potentially involving migratory birds as well as other ecological and anthropogenic drivers [21]. However, the emergence of geographically structured clades and evidence of sustained local transmission indicated that WNV can establish persistent transmission cycles following introduction. The increasing dominance and broader connectivity of lineage 2 further support its adaptive success within the European ecological context, in line with its documented expansion and replacement of lineage 1 in several regions [22]. Importantly, recent genomic evidence also indicates that the contemporary expansion of WNV Lineage 2 is no longer restricted to northern Italy. Although only a limited number of genomes from central and southern Italy were available for inclusion in the present analyses, these sequences cluster with the Balkan Lineage 2 diversity, suggesting an independent introduction into southern Italy during the most recent transmission seasons. This pattern, together with the marked increase in reported cases during 2025, supports the need for intensified genomic surveillance in central and southern regions [8, 23].

Our study also revealed historical mismatches between epidemiological and genomic surveillance representativeness. At a national level, sequencing efforts broadly tracked human and mosquito case records, which was not the case for other vertebrate hosts. Regionally, the Northern macroregions were well represented in both epidemiological and genomic surveillance, while all other regions showed imbalances between these two types of surveillance or generally low surveillance across host types. Such imbalances limit the capacity to accurately reconstruct transmission pathways, detect emerging variants and hence fully characterise multi-host dynamics. This aligns with broader concerns in genomic epidemiology, where uneven sampling remains a major constraint on inference [24], and highlights the need for more integrated One Health surveillance strategies that capture the full ecological complexity of WNV transmission.

Overall, the historical and novel data presented here point toward an eco-epidemiological system undergoing change, in which climatic conditions and other properties of host and vector ecological suitability are jointly driving the expansion and persistence of WNV in Italy. The convergence of epidemiological, environmental and genomic signals underscores the need for integrated surveillance frameworks capable of capturing these multidimensional processes. In the context of ongoing environmental change, such integration will be essential not only for understanding current transmission dynamics, but for anticipating future risk and guiding targeted public health interventions in Italy and elsewhere.

## Limitations

Several limitations should be considered when interpreting the data and modelling outputs presented. Historical surveillance efforts in Italy progressively evolved over the study period, reflecting the transition towards an increasingly integrated One Health surveillance framework while remaining spatially and temporally heterogeneous due to its risk-based implementation across regions. Accordingly, long-term observed and modelled epidemiological trends should be interpreted in light of changes in surveillance intensity and reporting practices that may have influenced the historical detection of cases. Although sequencing broadly tracked human and mosquito surveillance at the national level, regional mismatches—particularly for vertebrate hosts—may have obscured intermediate transmission links and limited the resolution of inferred dispersal pathways and multi-host transmission dynamics. In addition, the relatively limited availability of host-derived genomes constrains fine-scale reconstruction of transmission processes, meaning that phylogeographic patterns should be interpreted as broad indicators of historical connectivity and circulation rather than fully resolved transmission networks. Our analyses also do not explicitly account for many relevant factors, such as ecological processes within reservoir host populations, with bird abundance, movement patterns, species-specific competence and ecological interaction, and population immunity strongly modulate mechanisms underlying WNV persistence and spread. Furthermore, environmental variables were analysed at aggregated spatiotemporal scales, and the use of linear modelling approaches may underrepresent local ecological heterogeneity and non-linear environmental and eco-epidemiological effects. Despite these limitations, the integration of genomic, epidemiological, and environmental data provides one of the most comprehensive reconstructions of WNV evolutionary, ecological, environmental and spatio-temporal dynamics currently available for Italy, highlighting the value of integrated One Health surveillance frameworks for understanding and anticipating future transmission risk.

## Methods

### Evidence of circulation review

A literature review following PRISMA guidelines was conducted using searches in PUBMED and SCOPUS (last accessed: 30/12/2025), complemented by sources reported in the review study of Abbas et al. [1] (**Figure S1**). The PUBMED query was: “((west nile) OR (west nile virus) OR (WNV) OR (seroprevalence) OR (seropositivity) OR (seropositive) OR (seronegative) OR (italy) OR (neutralization) OR (neutralizing)) AND Italy”, which identified 94 unique papers. The SCOPUS query was “TITLE-ABS-KEY (((west nile) OR (west nile virus) OR (WNV)) AND ((seroprevalence) OR (seropositivity) OR (seropositive) OR (seronegative) OR (neutralization) OR (neutralizing)) AND Italy)”, which identified in 70 unique papers. The two queries, complemented with the Abbas et al. study, identified 50 unique papers with evidence of WNV past circulation in Italy (**Figures S1-3**). Studies were eligible for inclusion if they reported evidence of WNV circulation derived from serology (e.g., seroprevalence, neutralisation assays), molecular detection (PCR-based assays) or genomic sequencing data across vertebrate or vector hosts. Experimental studies and reports lacking primary data were excluded. Data were extracted from the 50 unique papers, resulting in a total of 169 data points. For simplicity: reported hosts were aggregated into four categories - human, equine, bird and other vertebrates; when longitude-latitude coordinates were not reported the centroid coordinates of the location (e.g., town, city), commune or region (official administrative boundaries) were used; and we visually present prevalence and positive rates on the same quantitative scale. **Table S1** includes details of the reviewed papers.

### Climate data

Monthly climate data was obtained from <u>Copernicus.eu</u> for the period 1979 to 2025, including three variables: temperature, precipitation, and humidity. The specific satellite data set “Essential climate variables for assessment of climate variability from 1979 to present” was used. The data was extracted per macroregion using the R-package *Terra* and summarized as yearly means (**Figure S7**).

### Time series trends

Long-term trends in climatic or epidemiological variables were calculated using the Mann-Kendall test (for p-value) and Sen’s slope (for trend magnitude).

### Epidemiological data

Reported cases were obtained from the Italian Ministry of Health through the national WNV surveillance system (EpiInfo platform), as reported by the Istituto Superiore di Sanità (ISS). Cases were stratified by host type (human, equine, mosquito, and bird) and subsequently aggregated by macroregion (**Figure S4**). Human cases were defined according to national surveillance criteria and confirmed by laboratory testing, including detection of WNV-specific IgM antibodies and/or viral RNA by PCR.

### Relationship between climatic variables and epidemiological data

Linear regression models were applied to assess the relationship between the annual number of reported human cases and yearly mean climatic variables at both the nationwide and macroregion spatial scales (**Figures 2, S8-9**). Since precipitation did not exhibit a statistically significant long-term trend (1979-2025), linear models were applied only to temperature and humidity. For each linear model, the correlation coefficient (Pearson’s) and p-value were reported.

### Age trends in epidemiological data

Reported cases were officially aggregated into age groups: <=14, 15-44, 45-64, 65-74, >=75 (**Figures 1, S6**). The age trends among macroregions were statistically similar: homogeneous by Pearson’s Chi-squared Test (p-value=0.39), with no differences in slopes (cases by age) by likelihood ratio test (p-value=0.83) between the two GLMs, with and without the macroregion interaction term. To estimate the mean age of reported cases, midpoint values of each age group were used under two methods: (i) weighted mean by reported cases with analytical standard error, and (ii) bootstrap mean and respective 95% quantile. These provided comparable outputs (**Figure S6**). Using the 15-44 age group as reference (to avoid zero counts in some macroregions), age-specific risk ratios (for case reporting) per macroregion were estimated via the coefficients of a Poisson GLM (**Figure S6**).

### GLM modelling of human case records with time

Annual human case records (see *Epidemiological data*) were modelled at the macroregional level using climate variables (see *Climate data*) within a Generalized Linear Model (GLM) framework. Models were implemented using both the base stats package and the MASS package (v.7.3.65) in R (v4.5.3). Human case records were the response variable modelled, since it was the only host time series that exhibited a temporal trend consistent with long-term, climate-driven dynamics. The climate variables included temperature (T), relative humidity (H) and precipitation (P). Input data were aggregated annually per macroregion: for case records the sum was used; for climatic variables the spatial mean was used.

Poisson GLMs were initially fitted with climate variables as predictors and case counts as the response. Overdispersion diagnostics were assessed in two ways: the residual deviance divided by the degrees of freedom (deviance/df) and the sum of squared Pearson residuals divided by the degrees of freedom (Pearson χ^2^/df). Values were substantially greater than 1 for the Poisson GLMs, thus showing overdispersion (i.e., observed variance in case records exceeds that assumed by the Poisson distribution).

Negative Binomial (NB) GLMs were therefore adopted for all subsequent modelling (**Figure S10**). The inclusion of macroregion as a fixed effect was evaluated by a likelihood ratio test against an nbGLM without the fixed effect. The output LR=36 (p-value<0.001) supported the inclusion of the macroregion mixed effect. The final model took the form: *cases ~ T + H + R + macroregion* with the macroregion Centre as reference. Its summary output was:

| Predictor | Estimate | p-value | Sig. |
| --- | --- | --- | --- |
| Intercept (Macroregion: Centre) | -66.287 | $5.93 \times 10^{-6}$ | *** |
| Temperature (T) | 2.650 | $4.15 \times 10^{-8}$ | *** |
| Humidity (H) | 0.447 | 0.034 | * |
| Precipitation (R) | -1594.038 | 0.039 | * |
| Macroregion: Islands | -9.746 | $2.44 \times 10^{-10}$ | *** |
| Macroregion: North-East | 12.946 | $8.59 \times 10^{-14}$ | *** |
| Macroregion: North-West | 16.214 | $2.29 \times 10^{-12}$ | *** |
| Macroregion: South | -3.437 | $4.89 \times 10^{-5}$ | *** |

Significance codes: \*\*\* $p < 0.001$ , \* $p < 0.05$ . Reference macroregion: Centre.

Model fit was assessed via residual deviance, Akaike Information Criterion (AIC) and Pearson correlation between observed and fitted values (i.e., case records) per macroregion. Nationwide model performance was further evaluated by aggregating (i.e., summing) the fitted values across the five macroregions and computing root mean squared error (RMSE) and mean absolute error (MAE) against observed national yearly totals. Partial effects of climatic predictors were extracted and visualised using the visreg R-package (v2.8.0). A hindcast (1979-2012) was produced with the NBGLM over the full-time extent of the climate data set (see *Climate data*).

### Phylogenetic and phylodynamic analysis of viral spread

To investigate the genomic epidemiology of WNV in Italy, complete viral genome sequences (>10 kb) were retrieved from the NCBI Virus database (accessed 01 April 2026). Sequences were screened and assigned to lineages using the West Nile Virus Typing Tool implemented in Genome Detective. Lineage 1a (L1a) and lineage 2 (L2) sequences were retained for downstream analyses. After quality filtering and curation, the final data sets included 275 L1a sequences (92 from Italy) and 859 L2 sequences (328 from Italy). Sequences were aligned using MAFFT [25] and visually inspected in AliView [26]. Maximum likelihood phylogenies of these data were then inferred with IQ-TREE v.2 under the best-fit nucleotide substitution model [27]. The resulting phylogenetic trees were evaluated for temporal structure using the molecular clock framework in TreeTime, and sequences that did not conform to the expected temporal signal were excluded [28]. Time-calibrated phylogenies were then estimated using sampling dates (tip-dating), applying lineage-specific mean substitution rates based on previous estimates reported for WNV lineages (β = 3.56 × 10^−4^ substitutions / site / year for L1a and β = 2.46 × 10^−4^ for L2) [29]. Phylogeographic patterns were investigated using the mugration model implemented in TreeTime, enabling the inference of viral movement across geographic locations [30]. Spatiotemporal diffusion patterns were further explored through a custom Python-based workflow integrating phylogenetic outputs with spatial metadata. Bayesian phylogenetic analyses were conducted on monophyletic groups of Italian sequences identified in both lineages. Accordingly, two clades per lineage were analysed: clade i.i (n=16) and clade iv (n=54) for L1a; clade ii (L1) (n=131) and ii (L2) (n=164) for L2. Temporal signal was assessed using TempEst [31] and time-scaled phylogenies were inferred in BEAST v1.10.5 under a relaxed molecular clock and Skygrid coalescent prior [32]. Model selection was performed using marginal likelihood estimation through path-sampling and stepping-stone approaches, which supported the selected demographic and molecular clock models [33]. To model spatial diffusion within Italy, a relaxed random walk diffusion model with a Cauchy distribution was applied [34], allowing branch-specific variation in dispersal rates. For each sequence, latitude and longitude were attributed to the Italian macroregion from which the diagnostic sample was obtained. Markov chain Monte Carlo (MCMC) analyses consisted of two independent runs of 100 million iterations each, with sampling every 10,000 steps. Convergence and mixing were evaluated in Tracer v1.7.1, confirming effective sample sizes (ESS) >200 for all parameters. Maximum clade credibility trees were generated using TreeAnnotator after discarding the first 10% of samples as burn-in. We used the R package ‘seraphim’ [35] to extract and map spatiotemporal information embedded in the posterior trees.

To quantify lineage-specific spatial expansion dynamics, we utilized previously described approaches to estimate wavefront distance trajectories from posterior phylogeographic reconstructions [35]. For each posterior tree, the maximal wavefront distance (MWD) was calculated as the greatest geographic distance between the inferred epidemic origin and the spatial coordinates associated with dispersal events through time. Temporal variation in wavefront distance (VWD) was computed as the change in maximal wavefront distance between consecutive temporal intervals, allowing quantification of dispersal jump magnitude during lineage expansion. Wavefront statistics were summarized across posterior trees to obtain median estimates and associated 95% highest posterior density (HPD) intervals.

To investigate the relationship between dispersal dynamics and environmental gradients, climatic variables associated with inferred source-sink locations and time points of dispersal jumps were extracted (see *Climatic data*) and stratified by season (**Figures S11-12**). Quantile regression analyses were performed to evaluate associations between variation in wavefront (distance of dispersal events) and concurrent variation in climatic variables (temperature, humidity, and precipitation) across the 5%, 50%, and 95% quantiles of dispersal variation (**Figure S13**).

## Data Availability

All data produced in the present work are contained in the manuscript

## Statements

### Data availability

NA

### Conflict of interest

None declared.

### Funding statement

This work was supported by the Novo Nordisk Foundation (NNF24OC0094346), by a National Health & Medical Research Council (Australia) Investigator grant (GNT2017197) to ECH, and by the South African Medical Research Council with funds received from the South African National Department of Health and the UKRI Medical Research Council, with funds received from the UK Government’s International Science Partnerships Fund (grant 96881).

### Ethical statement

NA

### Use of artificial intelligence tools

None declared.

## Acknowledgement

The authors acknowledge the public data repositories provided by ISS Epicentro and the ArboItaly platform.

## Notes

### Competing Interest Statement

The authors have declared no competing interest.

