## Appendix for "West Nile virus in Italy: history and evolving transmission patterns"

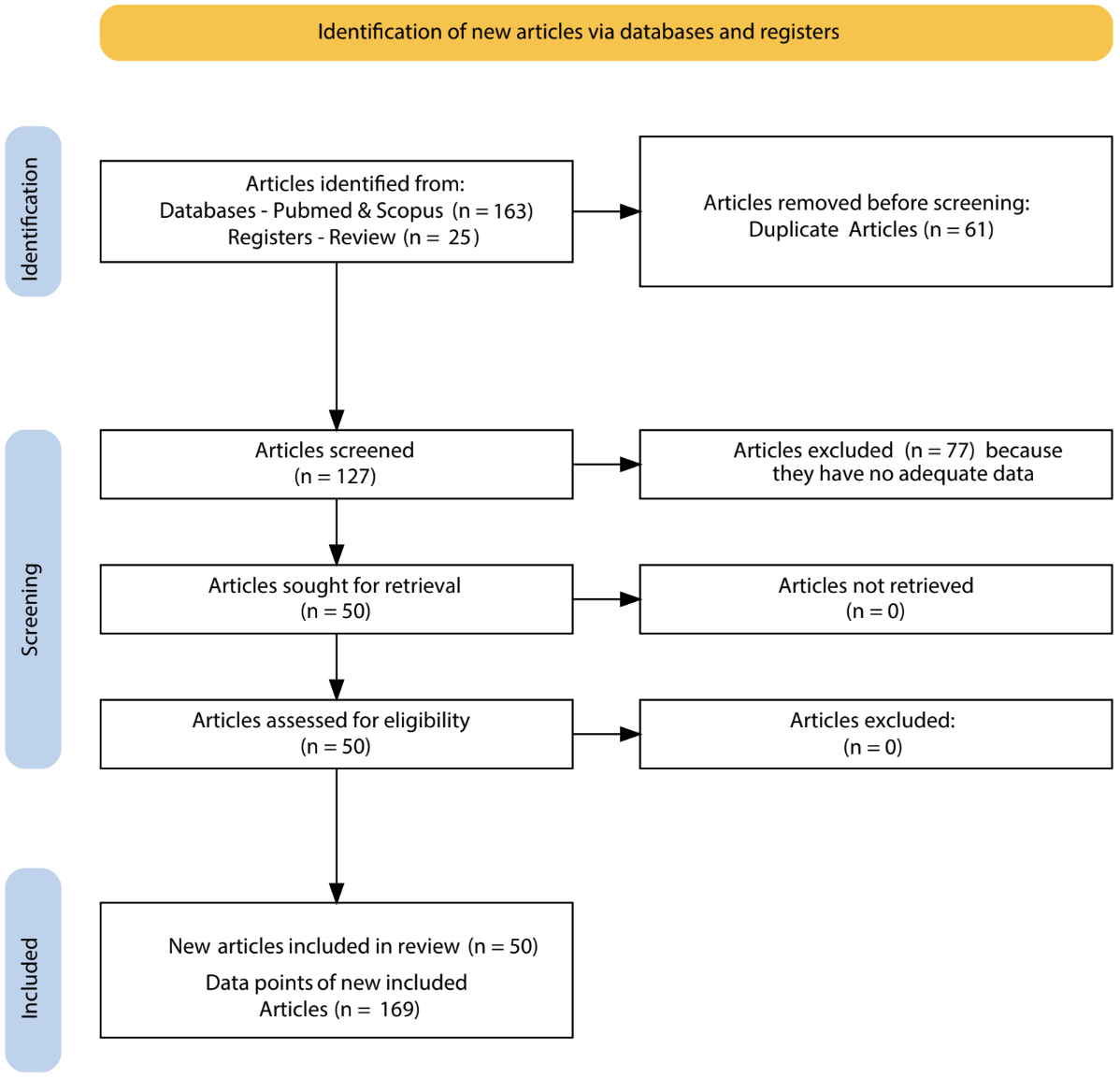

**Figure S1. PRISMA flow diagram of study selection.** Flowchart summarising the identification, screening, eligibility assessment, and inclusion of studies in the analysis. A total of 188 records were identified from databases (PubMed and Scopus) and registers, of which 61 duplicates were removed prior to screening. Following title and abstract screening (n = 127), 77 records were excluded due to insufficient or irrelevant data. Fifty full-text articles were assessed for eligibility, all of which were included in the final analysis. No records were excluded at the full-text stage. In total, 50 studies were included, contributing 169 data points.

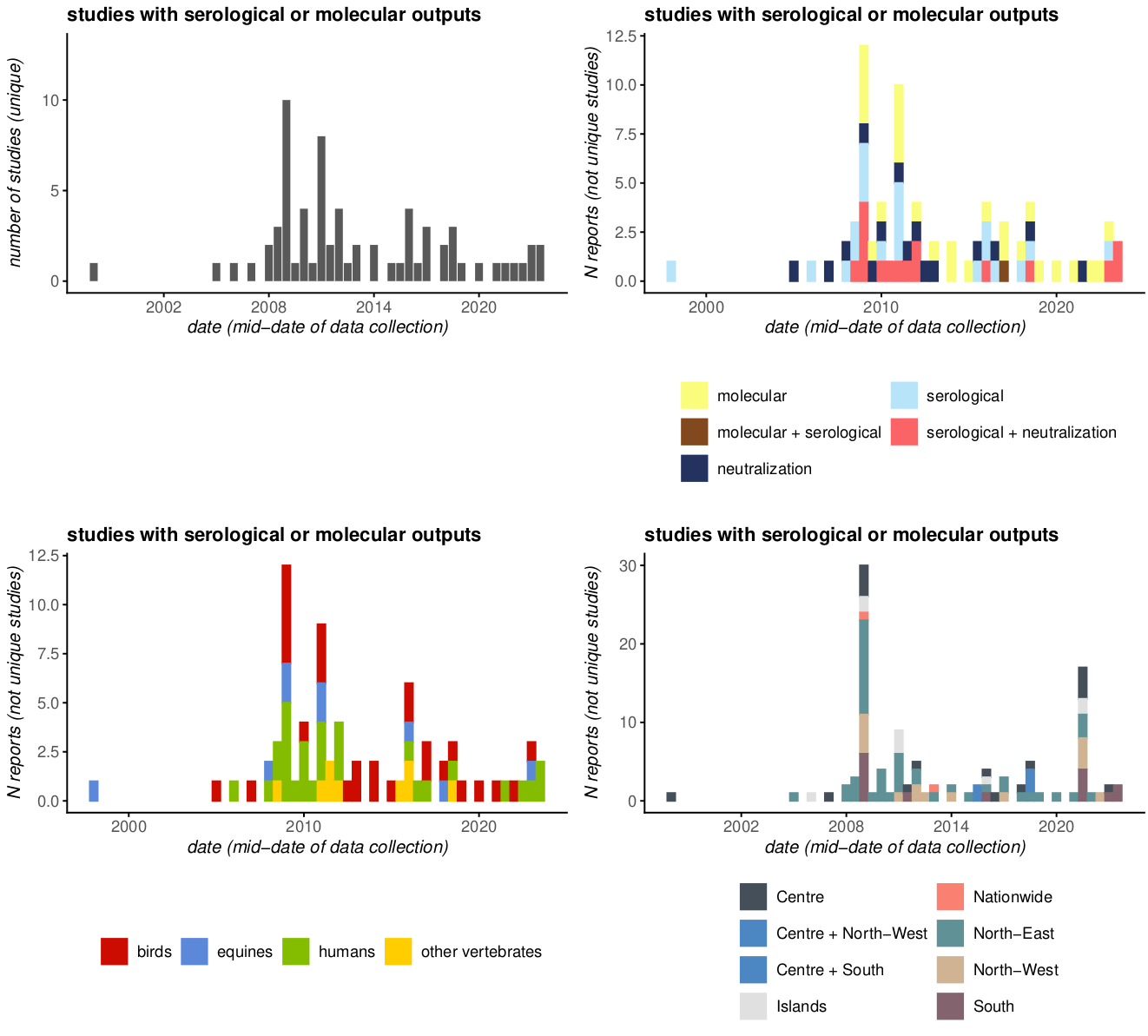

#### **Figure S2. Temporal distribution and characteristics of data records from the literature review.** Top-left) Yearly number of unique studies reporting serological or molecular evidence of WNV in Italy. The year is based on the midpoint date of data collection; Top-right) Yearly number of reported data points stratified by diagnostic approach; C) Yearly number of reported data points by host category; D) Yearly reported data points across Italian macroregions or nationwide (some records are reported with spatial ranges that span multiple macroregions).

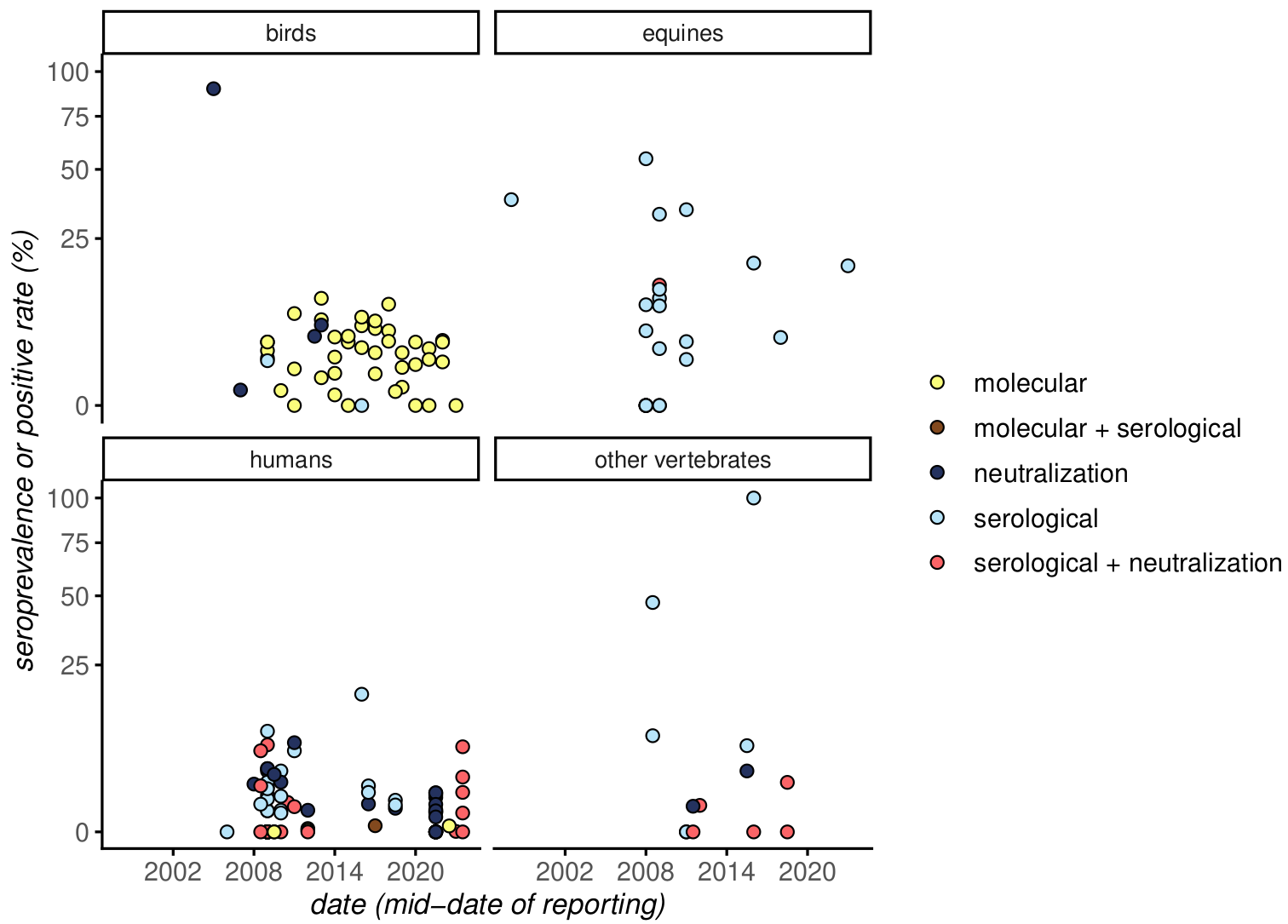

#### **Figure S3. Temporal distribution of data records from the literature review by diagnostic approach and host category.** Yearly distribution of reported seroprevalence or (molecular) positivity rate across host categories, where each point represents a reported estimate / measure from an individual study, plotted according to the year midpoint of the data collection period. Colours indicate the diagnostic approach (some data records include combinations of diagnostics to reach an estimate / measure).

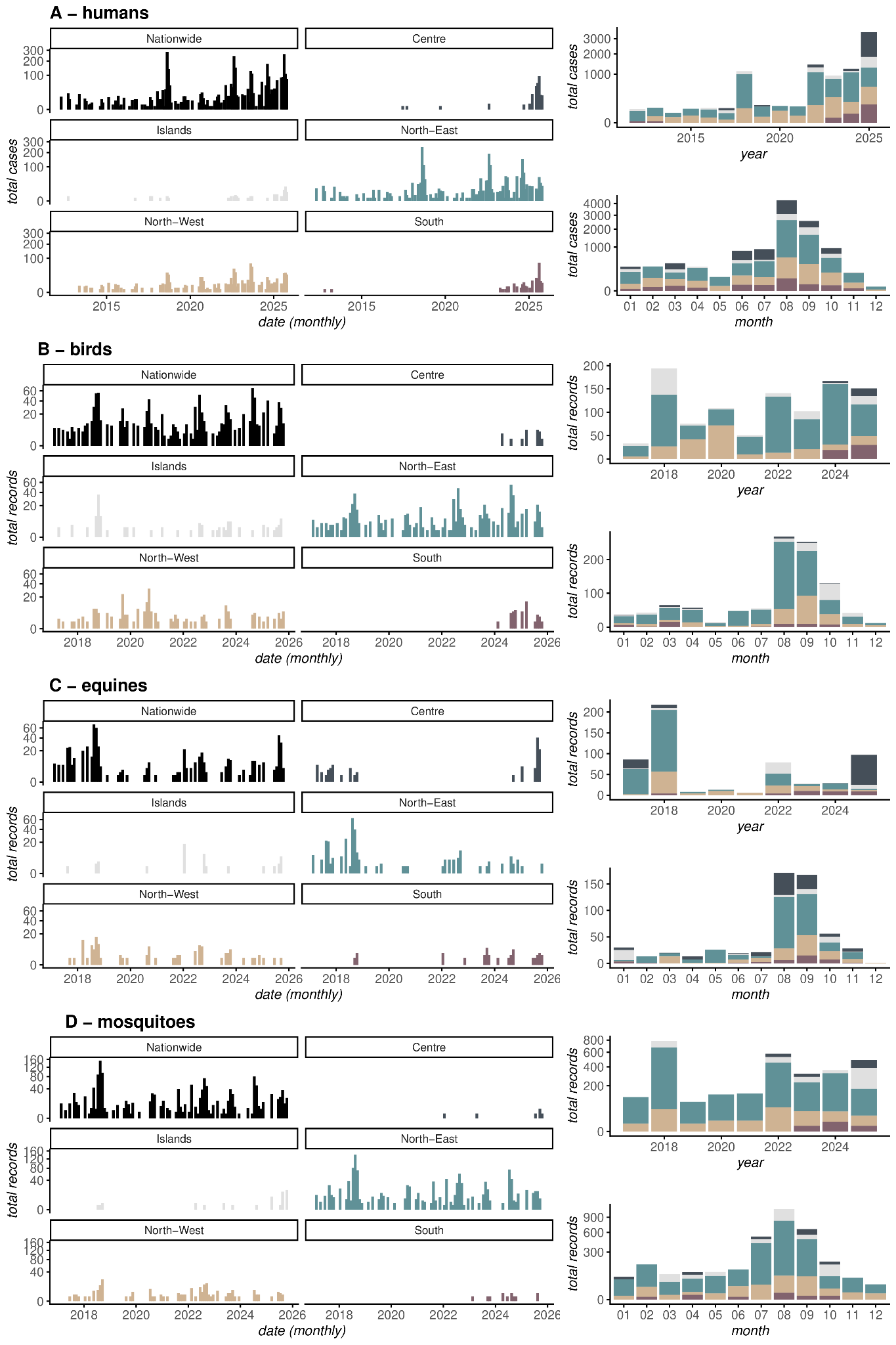

#### **Figure S4. Temporal dynamics of WNV case records across host categories and Italian macroregions.** A) humans, B) birds, C) equines and D) mosquitoes. The left column shows monthly counts of reported cases (humans) or records (animals and vectors) at the national level and across macroregions (Centre, Islands, North-East, North-West and South). The right column summarizes the same data as aggregated yearly sums (top) and across all years monthly sums (bottom), with colours indicating macroregions. For some of the subpanels the Y-axis has been square-root transformed to ease visualization.

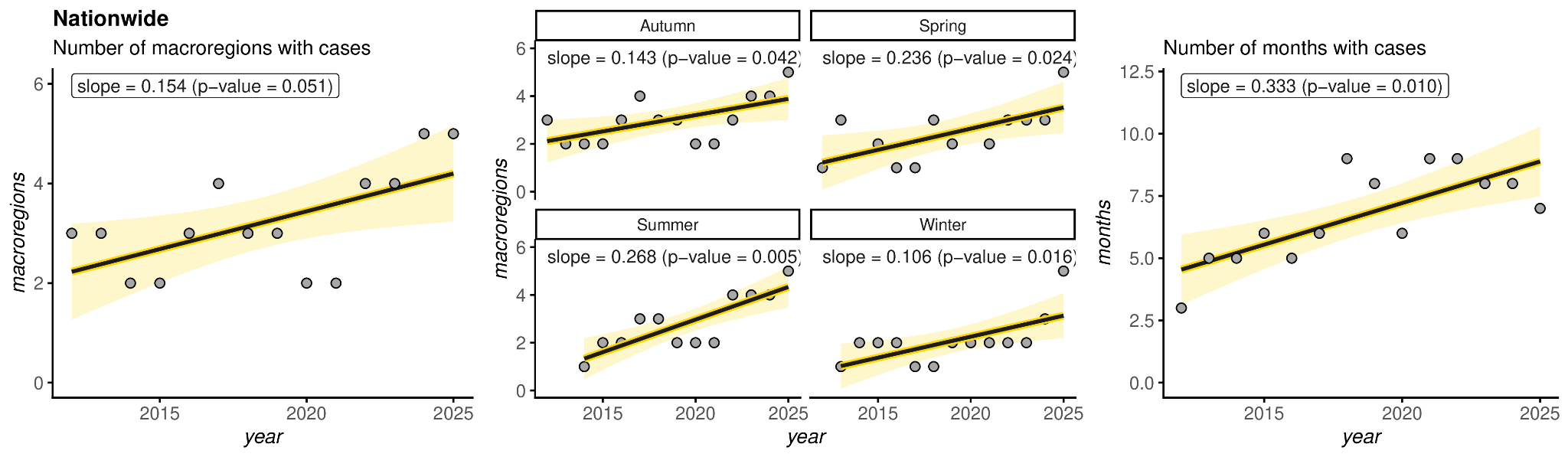

**Figure S5 - Spatio-temporal changes of non-zero human case records.** Left) Number of macroregions a year (among the five possible - North-West, North-East, Centre, South, Islands) that had non-zero human case records (i.e. cases >=1) over time; Center) Same as in the Left panel, but evaluated per season (Autumn = Sep-Nov, Spring = Mar-May, Summer = Jun-Aug, Winter = Dec-Feb); Number of months a year (among twelve possible) that had non-zero human case records (i.e. cases>=1) over time.

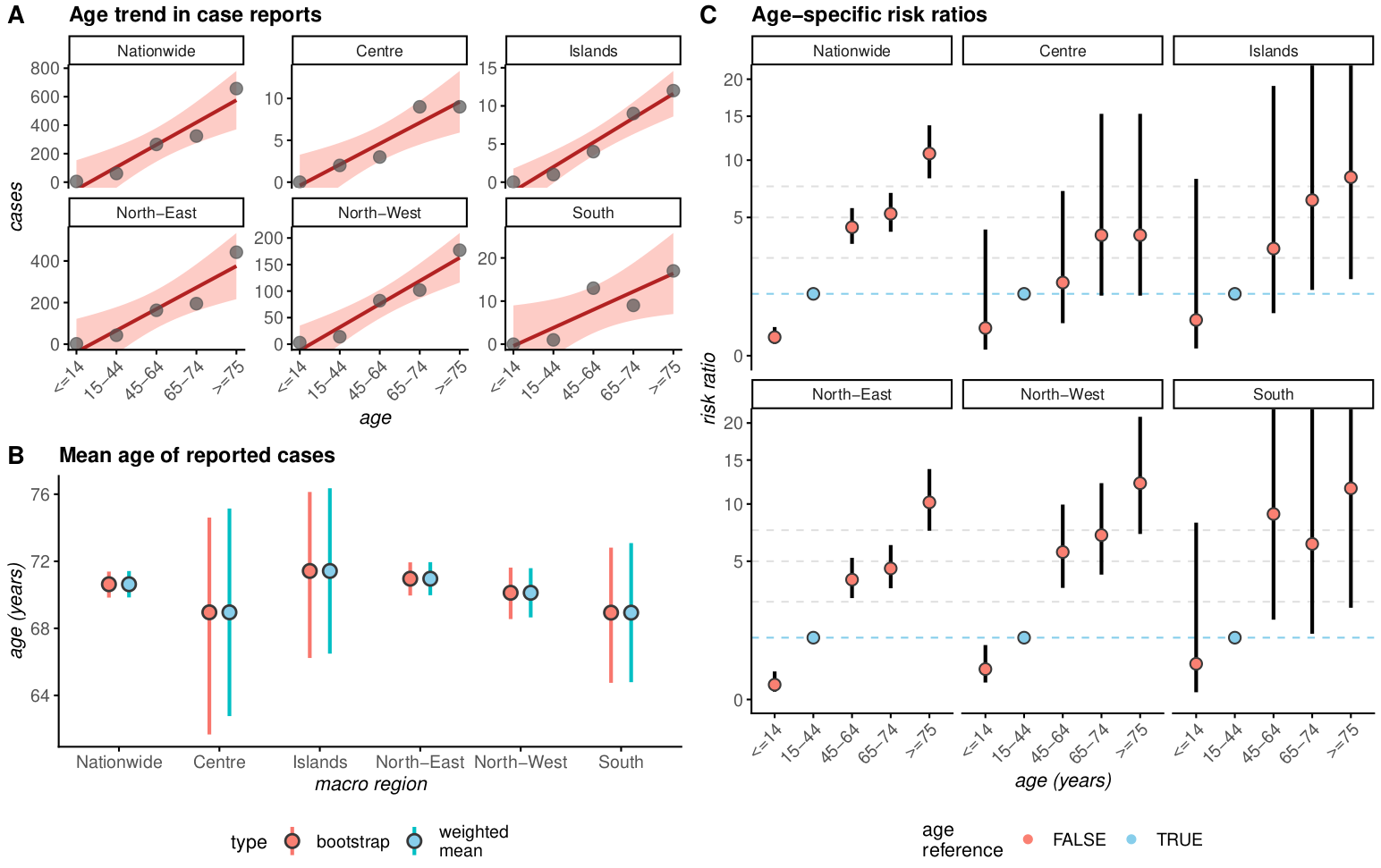

### **Figure S6. Age-related characteristics of WNV human case records across Italian macroregions.** A) Distribution of reported human cases by age group (sum of records) across Italy and macroregions for the entire observation period, with fitted linear trends; B) Estimated mean age of reported cases by macroregion, calculated using both weighted averages and bootstrap approaches (whiskers represent uncertainty estimates); C) Age-specific risk ratios (RR) for case reporting across macroregions, estimated using a Poisson generalized linear model. The age group 15–44 years is used as the reference category (in blue). Points represent estimated RR and whiskers indicate 95% confidence intervals.

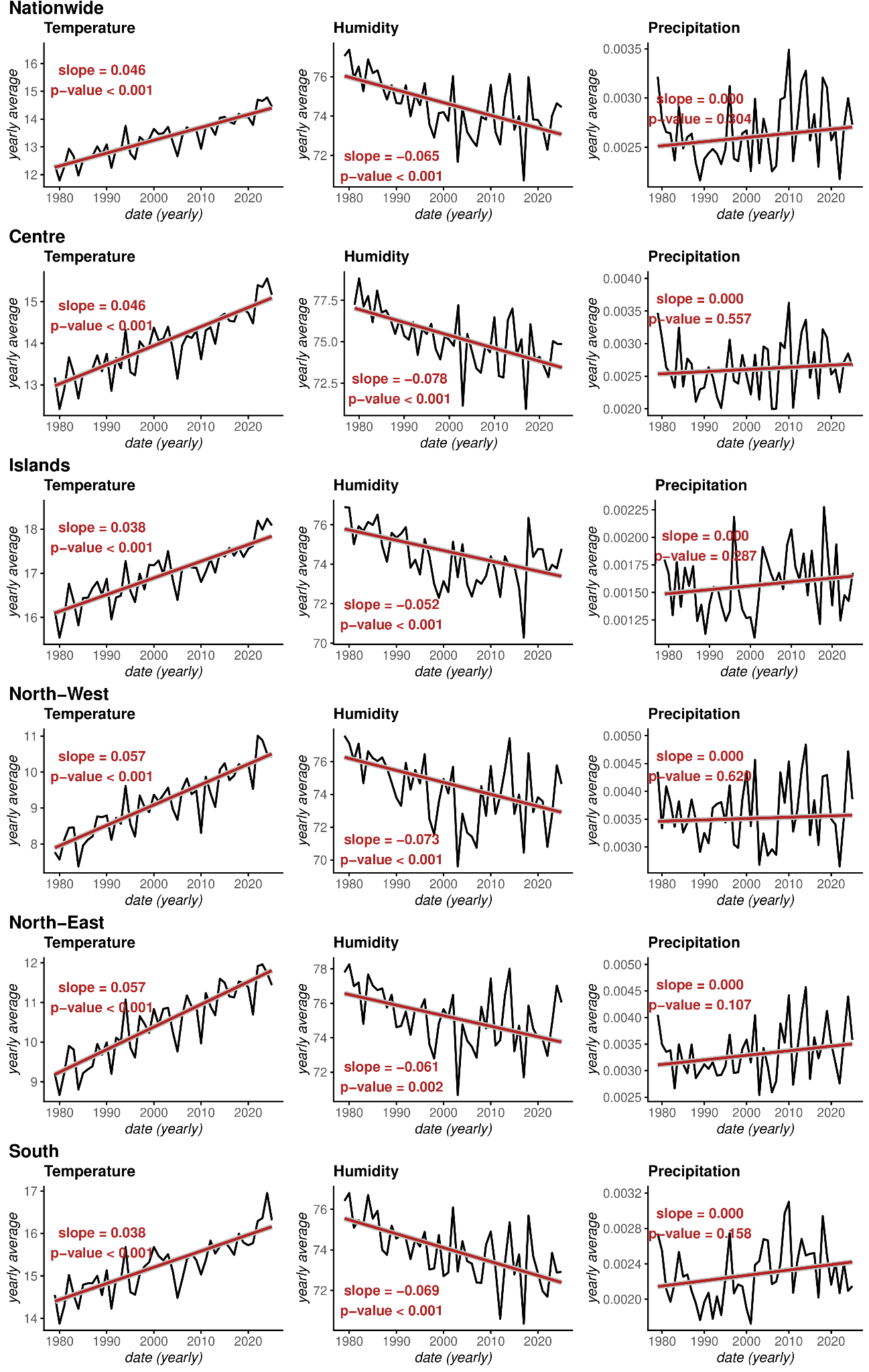

#### **Figure S7. Long-term climatic trends across Italy and macroregions.** Temporal trends in annual mean temperature, humidity and precipitation at the national level and across Italian macroregions (Centre, Islands, North-West, North-East and South). Each panel shows yearly values (black lines) and the corresponding linear trend estimated using Sen’s slope (red line). Slope coefficients and associated p-values (Mann–Kendall test) are reported within each panel.

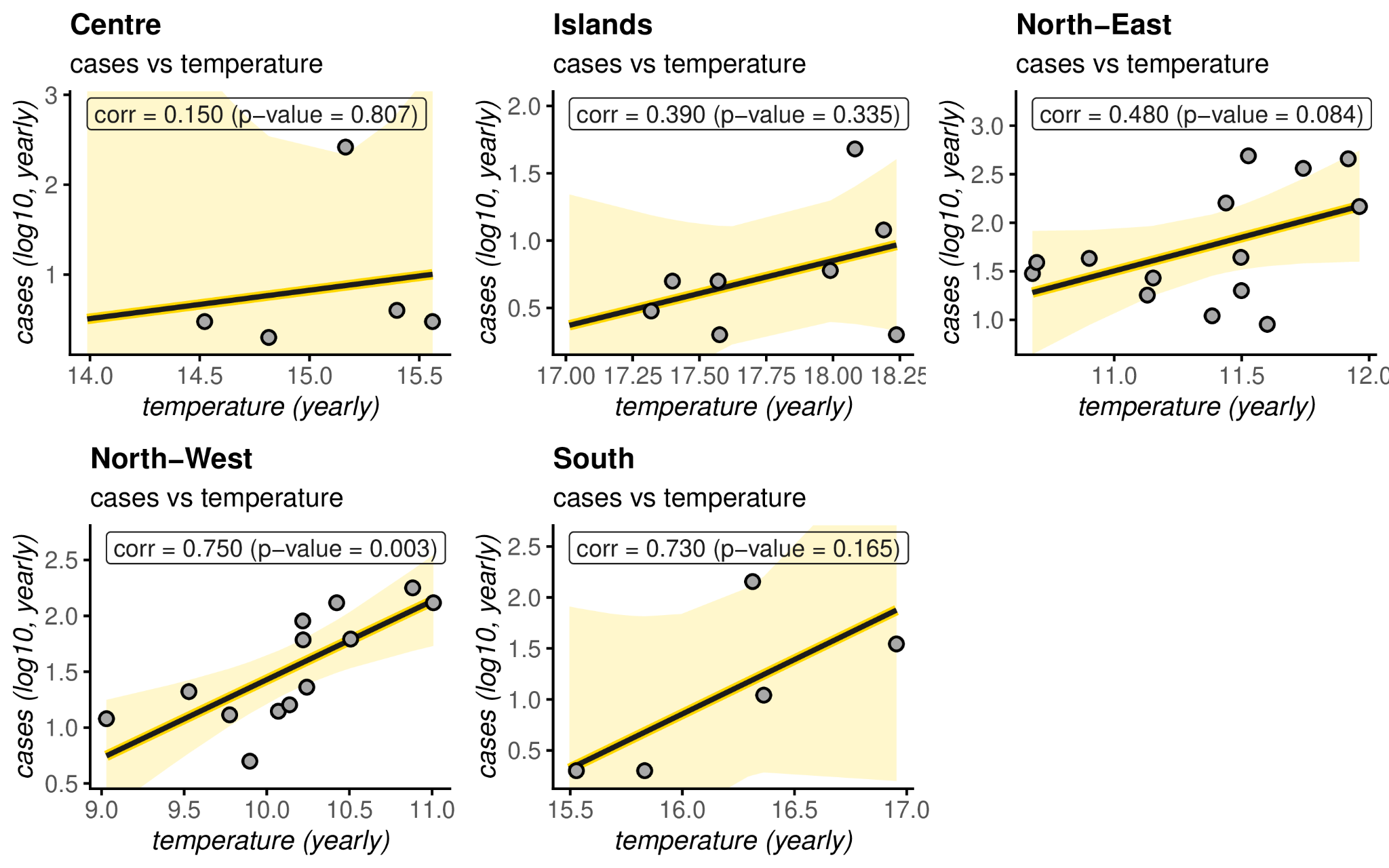

#### **Figure S8. Relationship between temperature and WNV human case records across Italian macroregions.** Each point represents yearly observations, with cases being the log10-transformed sum and temperature the geographic mean. Solid lines indicate fitted linear regression models, with shaded areas representing 95% confidence intervals. Pearson’s correlation coefficients (corr) and corresponding p-values are reported within each panel.

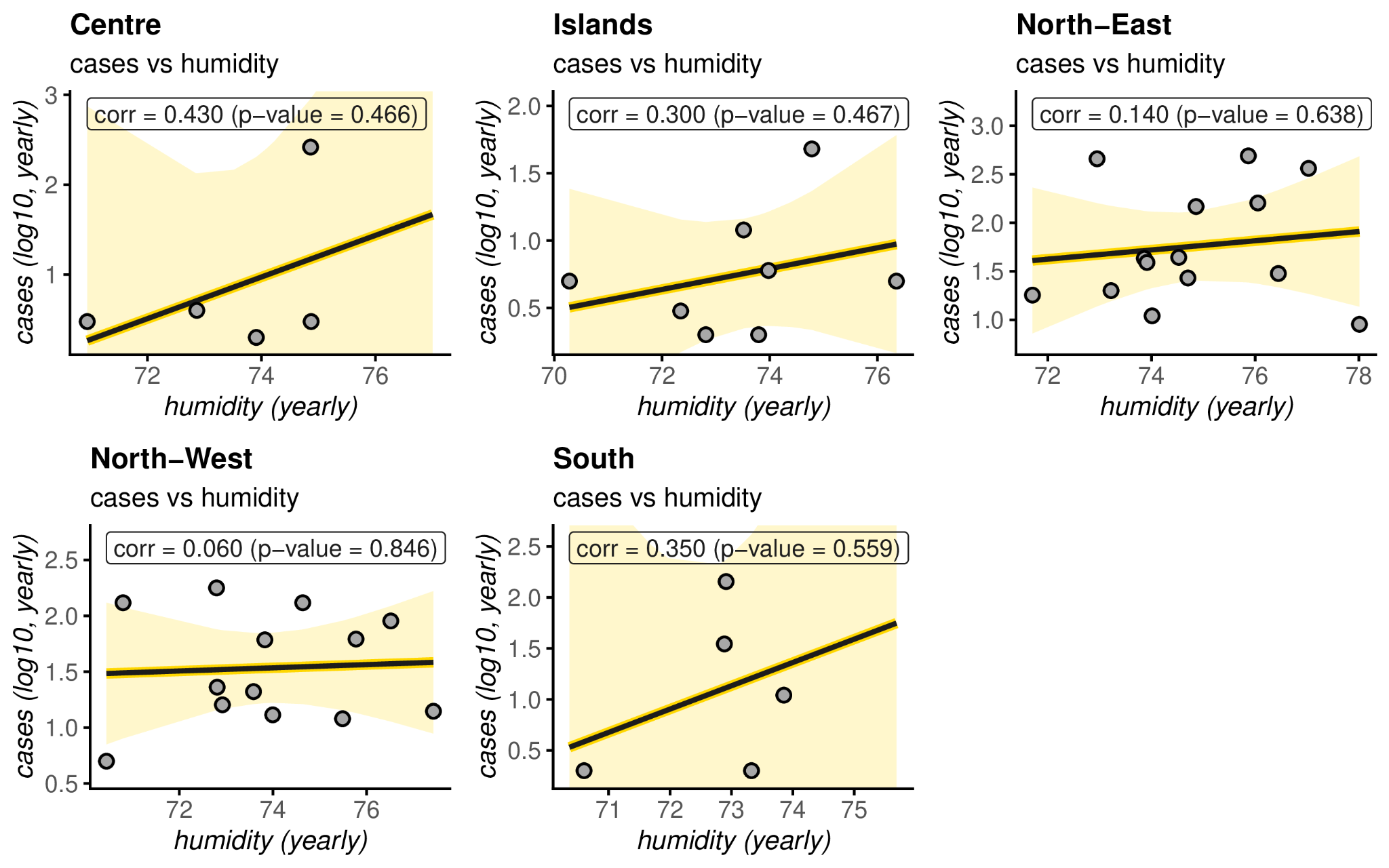

#### **Figure S9. Relationship between humidity and WNV human case records across Italian macroregions.** Each point represents yearly observations, with cases being the log10-transformed sum and humidity the geographic mean. Solid lines indicate fitted linear regression models, with shaded areas representing 95% confidence intervals. Pearson’s correlation coefficients (corr) and corresponding p-values are reported within each panel.

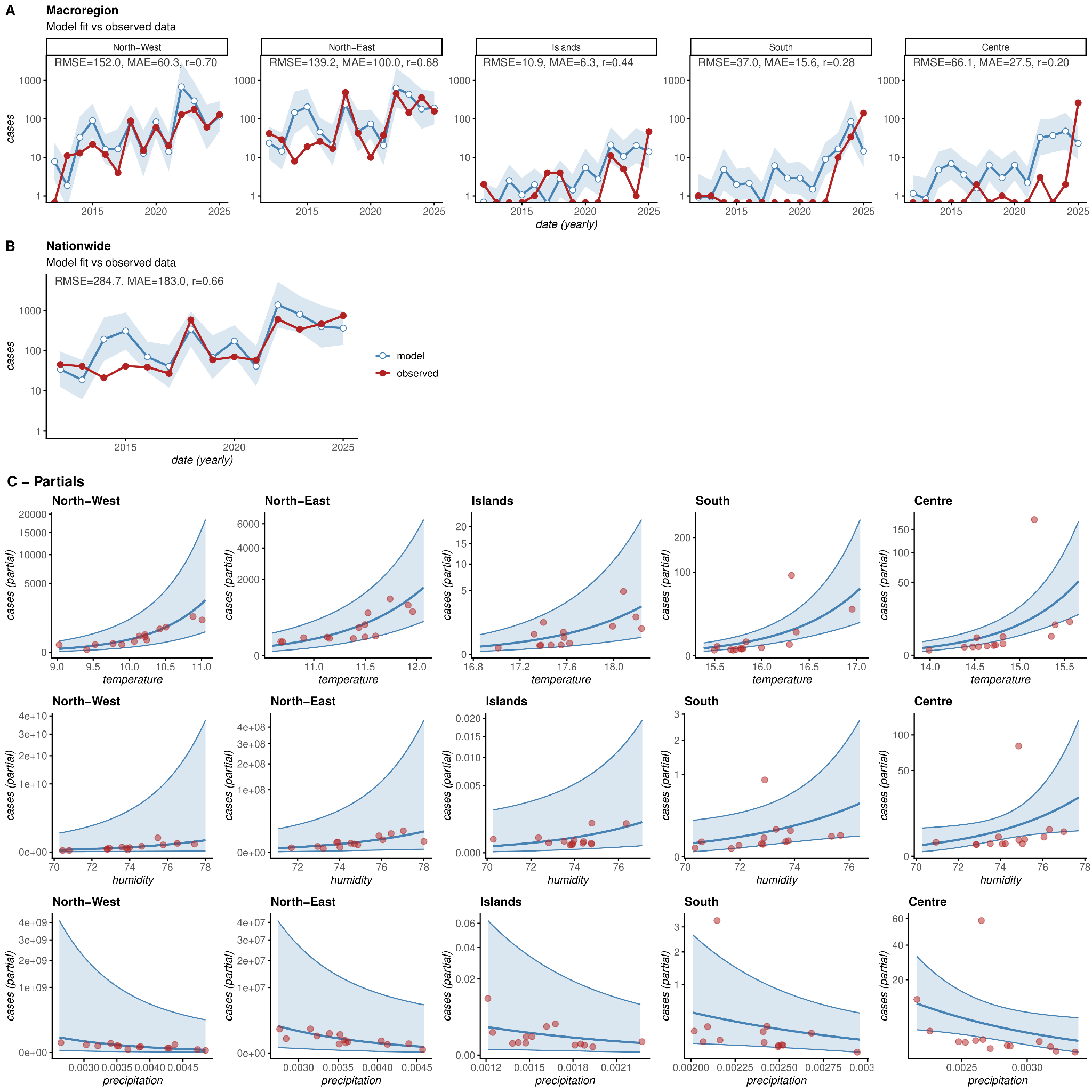

#### **Figure S10. GLM-related visualizations.** Panels show GLM outputs when fitting monthly WNV human case records with temperature, humidity, and precipitation as predictors, the macroregion Centre as reference and macroregion as a fixed effect. A) The resulting fits per macroregion, with inner labels presenting the root mean squared error (RMSE), mean absolute error (MAE) and Pearson’s correlation (r) (model in blue and data records in red). B) The resulting fit at the nationwide level (resulting from the sum of each macroregion’s fit as shown in Panel A), with inner labels presenting the root mean squared error (RMSE), mean absolute error (MAE) and Pearson’s correlation (r) (model in blue and data records in red). C) Partials per macroregion, per predictor variable (top = temperature, middle = humidity, bottom = precipitation).

**Supplementary Table S1.** Estimated times to the most recent common ancestor (tMRCA) and corresponding 95% highest posterior density (HPD) intervals for the major Italian WNV clades.

| ***Lineage*** | **Clade** | **tMRCA (95% HPD)** |
| --- | --- | --- |
| WNV L2 | ii_L2 | 2011.4 (2010.8–2012.3) |
| WNV L2 | II_11 | 2011.2 (2010.7–2011.8) |
| WNV L2 | iv | 2007.0 (2006.1–2008.0) |
| WNV L2 | v | 2024.8 (2024.0–2025.3) |
| WNV L2 | i | 2010.8 (2010.7–2011.5) |
| WNV L2 | iii | 2012.4 (2011.9–2013.5) |
| WNV L1a | iii | 2010.4 (2009.7–2011.0) |
| WNV L1a | ii | 2011.1 (2010.6–2011.5) |
| WNV L1a | iv | 2003.1 (2001.7–2004.7) |
| WNV L1a | i.i | 2006.3 (2005.6–2006.9) |
| WNV L1a | i.ii | 2010.1 (2009.6–2010.5) |
| WNV L1a | v | 2019.3 (2018.7–2019.8) |

tMRCA, time to the most recent common ancestor; HPD, highest posterior density interval

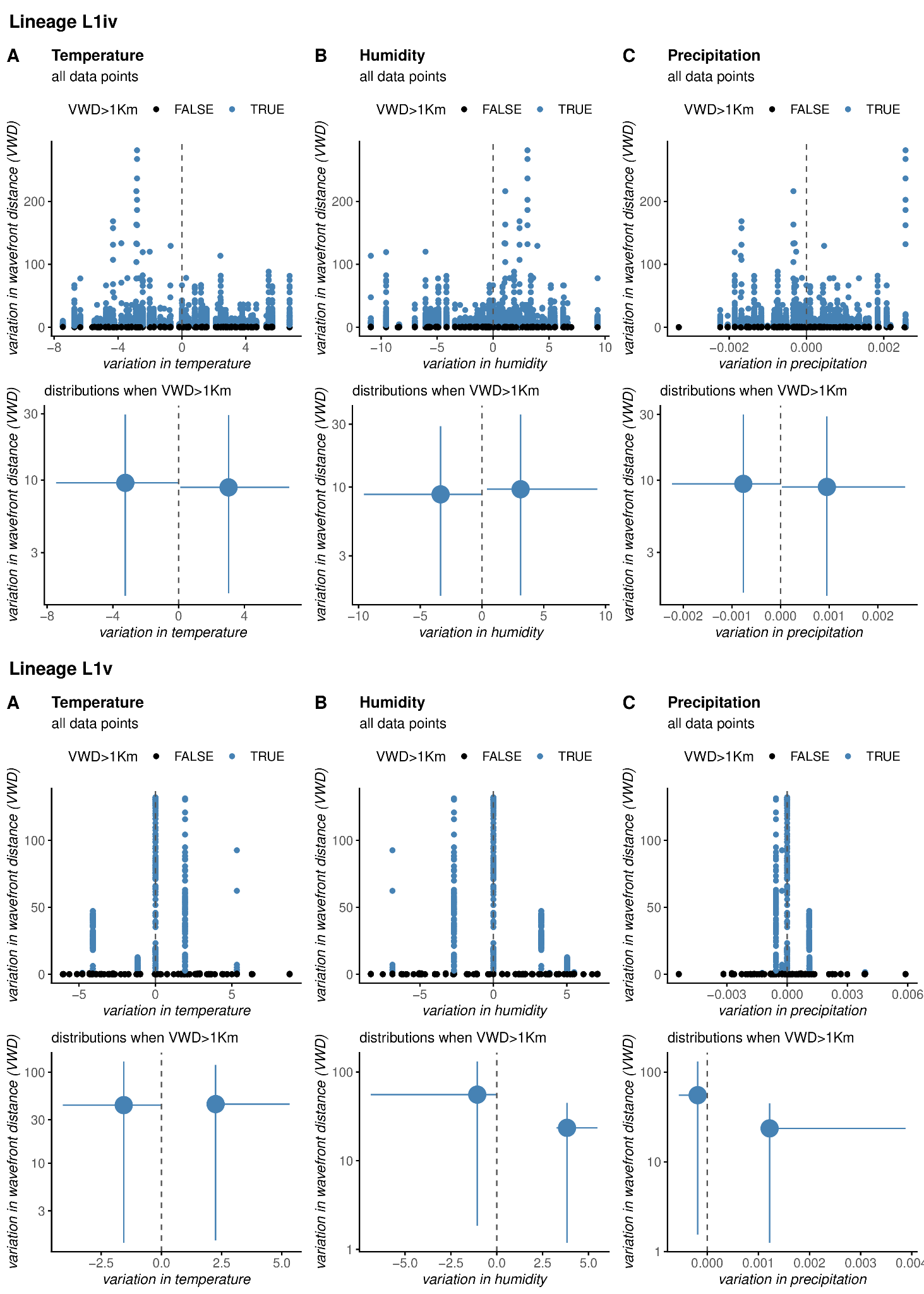

**Figure S11. Associations between variation in wavefront distance and climatic gradients across WNV lineages iv and v.** Relationships between variation in wavefront distance (VWD) and concurrent variation in climatic variables for WNV lineages L1iv and L1v. Panels show associations with temperature (A), humidity (B), and precipitation (C). Upper panels display all inferred dispersal events, with blue points indicating events associated with VWD >1 km and black points representing more localized dispersal events. Lower panels summarize the distribution of climatic variation associated with larger dispersal events (VWD >1 km). Vertical dashed lines indicate null climatic variation.

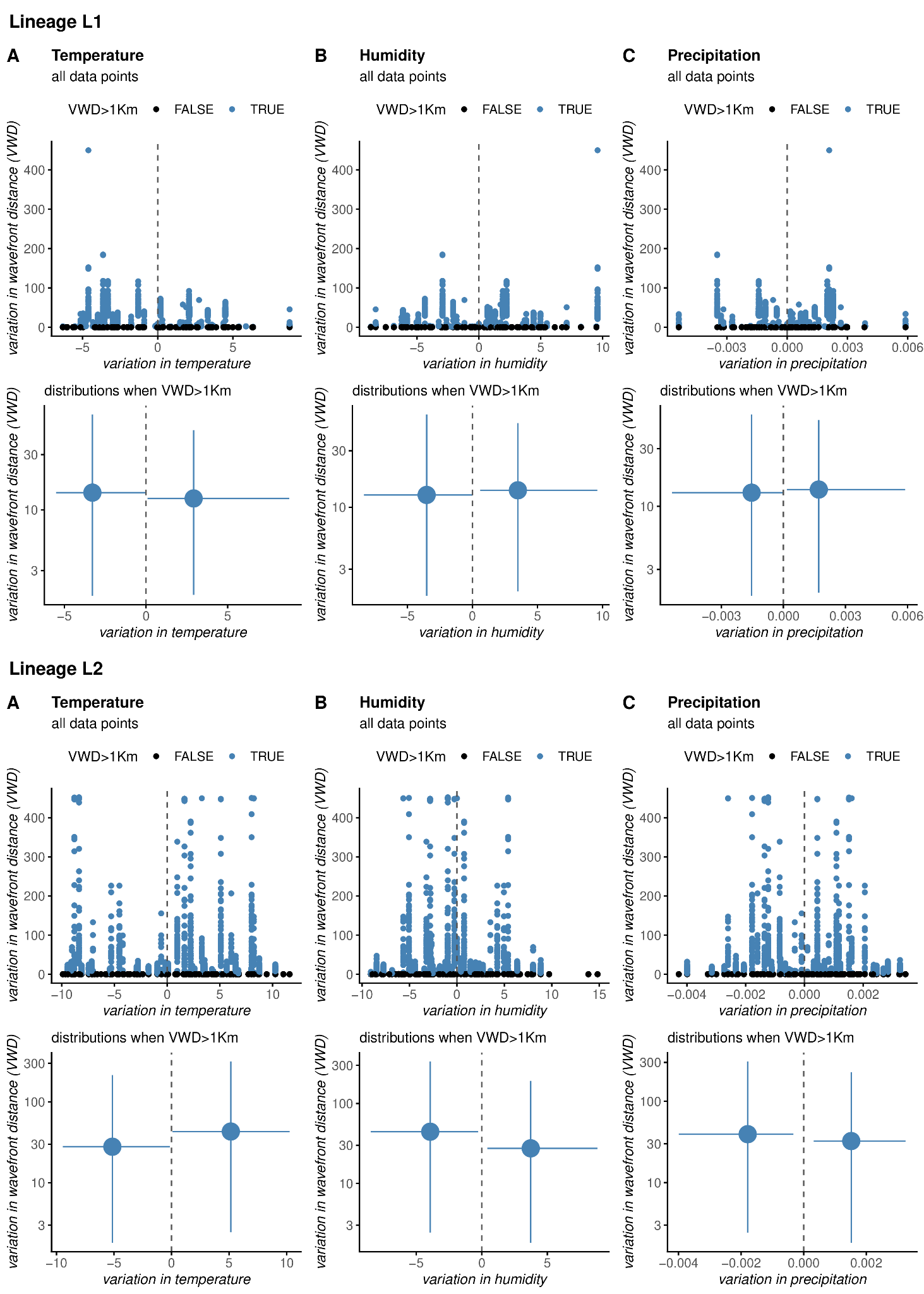

**S12. Lineage-independent associations between variation in wavefront distance and climatic gradients for WNV lineages L1 and L2.** Relationships between variation in wavefront distance (VWD) and concurrent variation in climatic variables for WNV lineages L1 and L2. Panels show associations with temperature (A), humidity (B), and precipitation (C). Upper panels display all inferred dispersal events, with blue points indicating events associated with VWD >1 km and black points representing more localized dispersal events. Lower panels summarize the distribution of climatic variation associated with larger dispersal events (VWD >1 km). Positive climatic variation values indicate movement toward regions with higher climatic values, whereas negative values indicate movement toward regions with lower climatic values. Vertical dashed lines indicate null climatic variation.

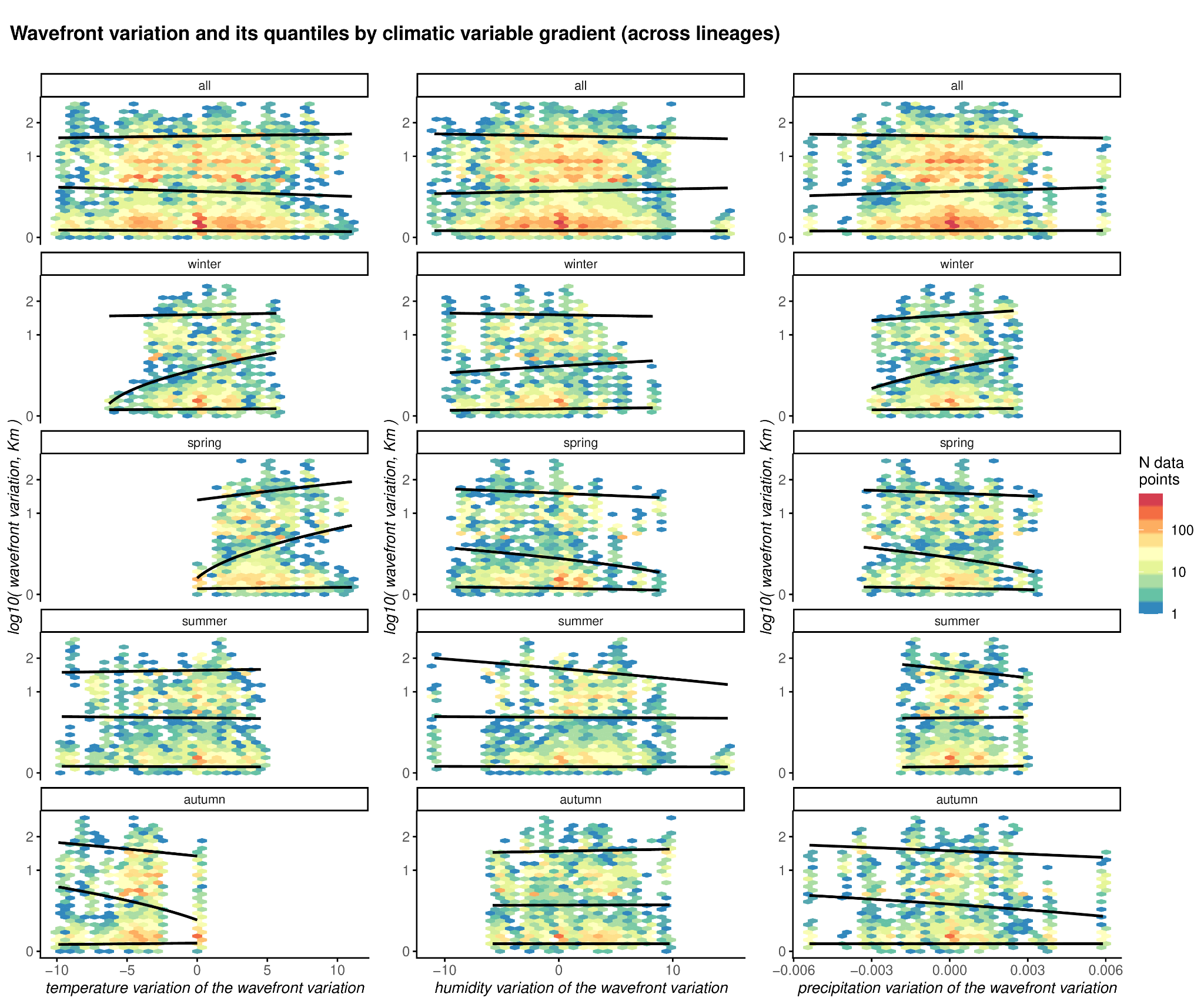

**S13**. **Quantile-specific associations between wavefront variation and climatic gradients across WNV lineages and seasons.** Density distributions showing the relationship between variation in wavefront distance and climatic gradients for temperature, humidity, and precipitation across all inferred dispersal events. Analyses are stratified by season (all, winter, spring, summer, and autumn). Black lines represent quantile regressions corresponding to the 5%, 50%, and 95% quantiles of wavefront variation. Color gradients indicate the density of observations (N data points). Positive climatic variation values indicate movement toward regions with higher climatic values, whereas negative values indicate movement toward regions with lower climatic values.

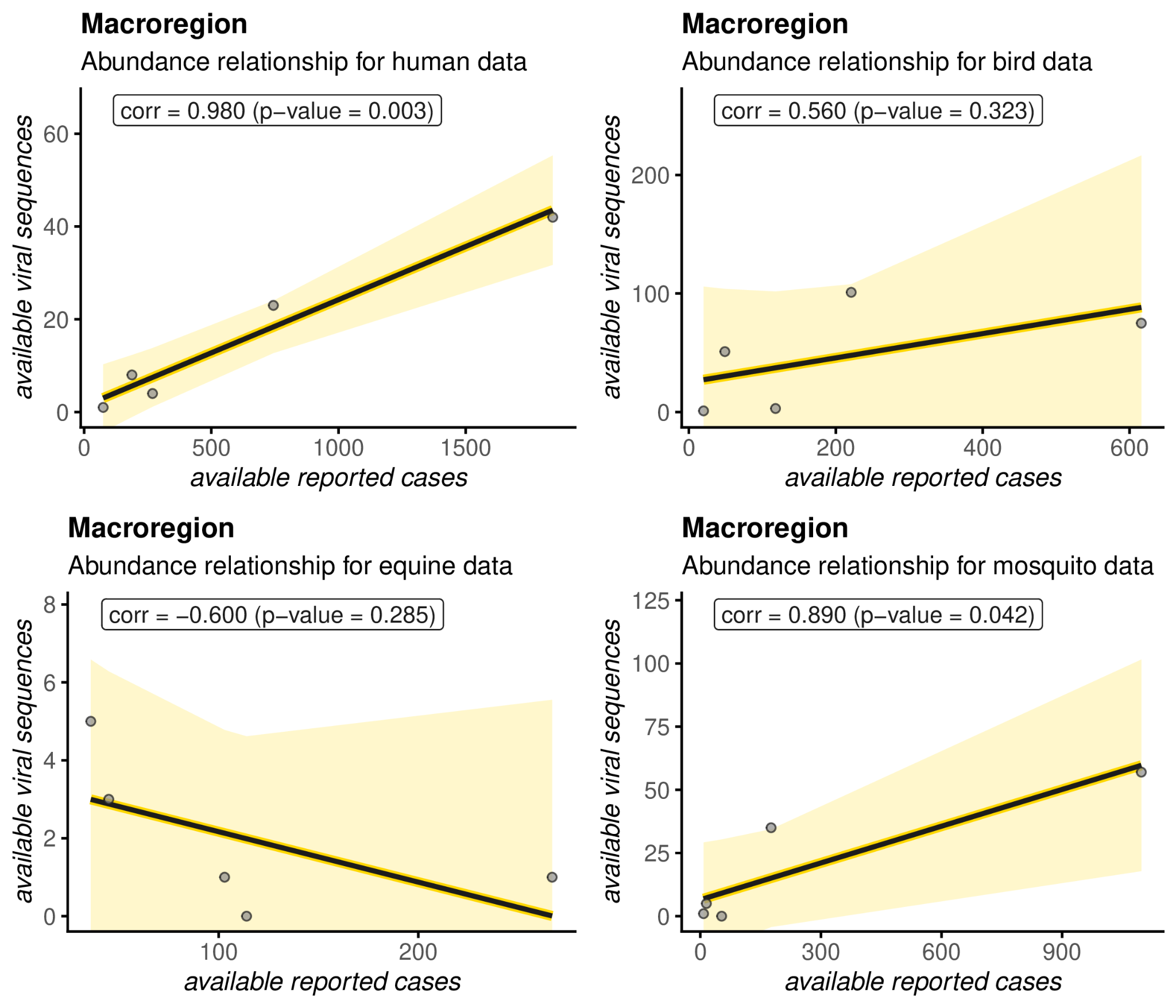

#### **Figure S14. Relationship of reported WNV case records versus available viral sequences.** Panels present scatterplots of the (abundance) relationship between case records and viral sequences per macroregion (grey points), and per host (different panels), with inner labels presenting Pearson’s correlation and respective p-value.

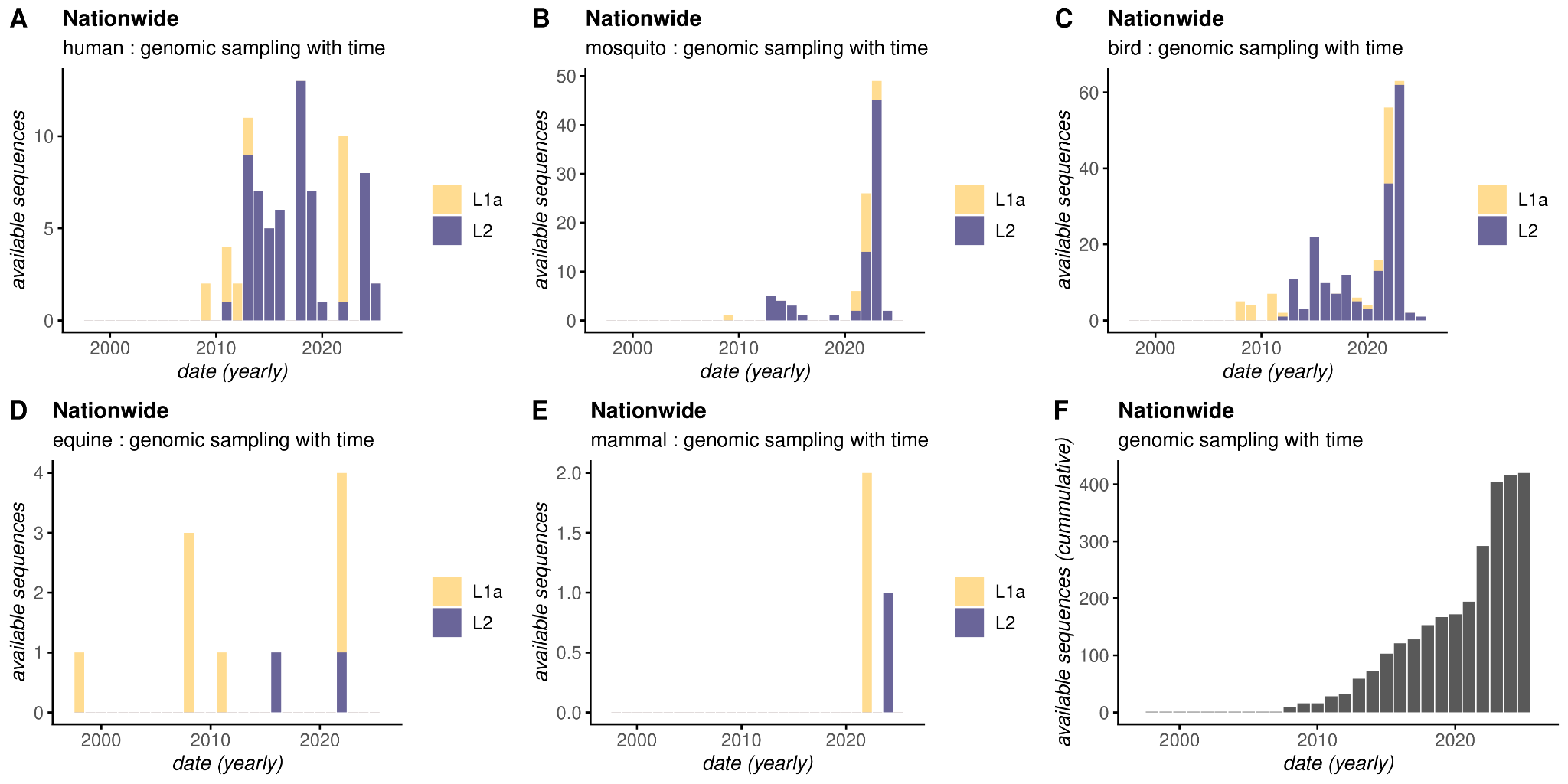

#### **Figure S15. Genomic sampling in time across Italy.** Panels A-E present the total number of available viral sequences per year, per host: A) Human, B) Mosquito, C) Bird, D) Equine, E) Mammal (non-specific). Totals by viral lineage are shown by color: yellow = L1A, purple = L2. F) Presents the (yearly) cumulative number of available viral sequences at the nationwide scale.

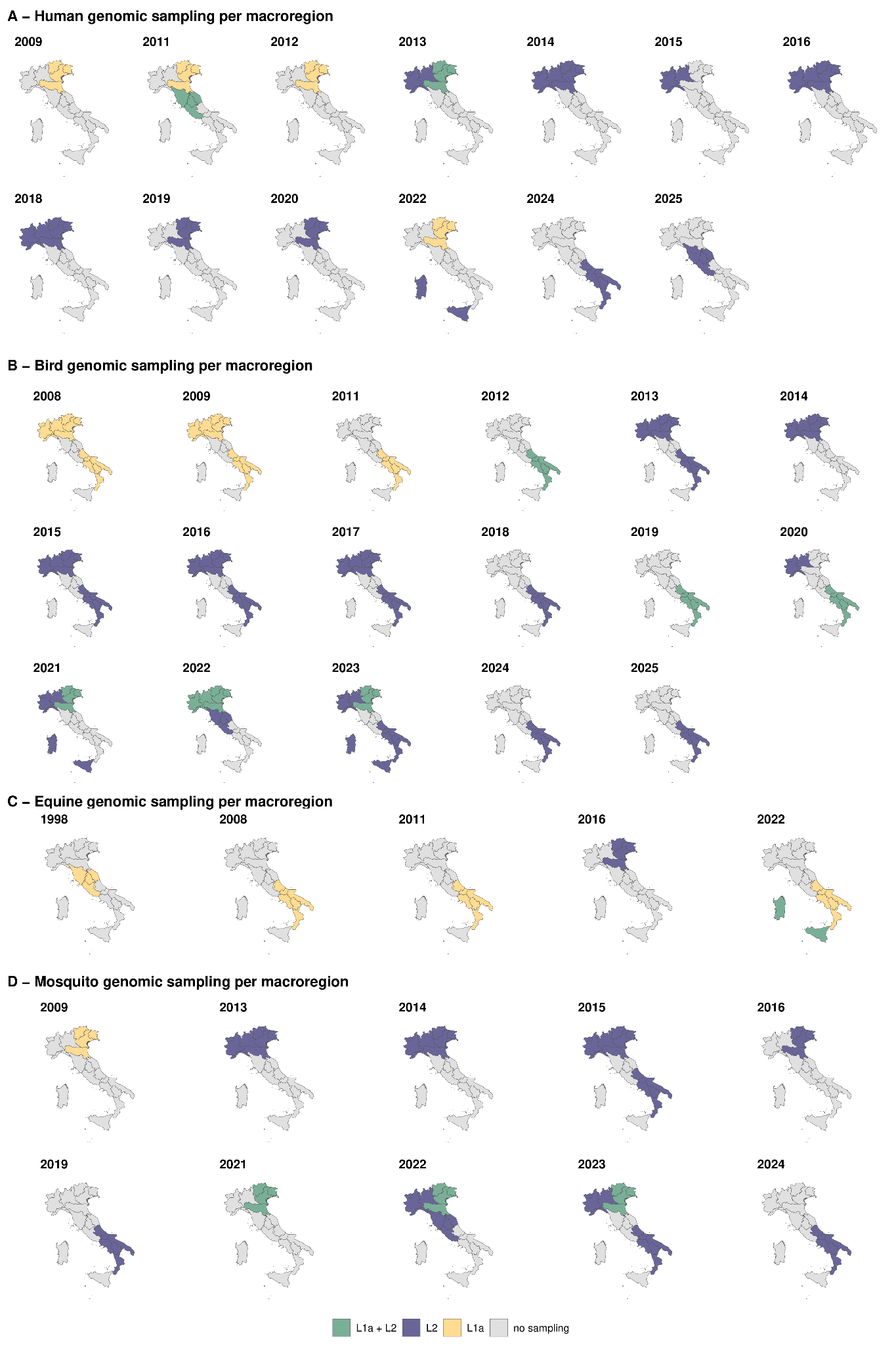

#### **Figure S16. Genomic sampling in time and space across Italy.** Panels A-D show the yearly macroregional presence status (i.e., whether at least one viral sequence was reported) of each viral lineage (both = green, yellow = L1A, purple = L2, grey = none) per host: A) Human, B) Bird, C) Equine, D) Mosquito. Years with no sampling are not presented (which vary per host type).
